# Multiplexed, quantitative serological profiling of COVID-19 from a drop of blood by a point-of-care test

**DOI:** 10.1101/2020.11.05.20226654

**Authors:** Jacob T. Heggestad, David S. Kinnamon, Lyra B. Olson, Jason Liu, Garrett Kelly, Simone A. Wall, Cassio M. Fontes, Daniel Y. Joh, Angus M. Hucknall, Carl Pieper, Ibtehaj A. Naqvi, Lingye Chen, Loretta G. Que, Thomas Oguin, Smita K. Nair, Bruce A. Sullenger, Christopher W. Woods, Gregory D. Sempowski, Bryan D. Kraft, Ashutosh Chilkoti

## Abstract

Highly sensitive, specific, and point-of-care (POC) serological assays are an essential tool to manage the COVID-19 pandemic. Here, we report on a microfluidic, multiplexed POC test that can profile the antibody response against multiple SARS-CoV-2 antigens—Spike S1 (S1), Nucleocapsid (N), and the receptor binding domain (RBD)—simultaneously from a 60 µL drop of blood, plasma, or serum. We assessed the levels of anti-SARS-CoV-2 antibodies in plasma samples from 19 individuals (at multiple time points) with COVID-19 that required admission to the intensive care unit and from 10 healthy individuals. This POC assay shows good concordance with a live virus microneutralization assay, achieved high sensitivity (100%) and specificity (100%), and successfully tracked the longitudinal evolution of the antibody response in infected individuals. We also demonstrated that we can detect a chemokine, IP-10, on the same chip, which may provide prognostic insight into patient outcomes. Because our test requires minimal user intervention and is read by a handheld detector, it can be globally deployed in the fight against COVID-19 by democratizing access to laboratory quality tests.

## Introduction

The ongoing severe acute respiratory syndrome–coronavirus 2 (SARS-CoV-2) pandemic poses an enormous challenge to the world. SARS-CoV-2 has resulted in over 47 million cases of coronavirus disease (COVID-19) worldwide, resulting in over 1.2 million deaths as of November 3, 2020 ^1^. Unlike many other viruses, SARS-CoV-2 displays high infectivity, a large proportion of asymptomatic carriers, and a long incubation time of up to 12 days, during which carriers are infectious ^2-4^. As a result, transmission has been widespread, resulting in overwhelmed healthcare capacities across the globe ^5,6^. Timely, reliable and accurate diagnostic and surveillance tests are necessary to control the current outbreak and to prevent future spikes in transmission.

Reverse transcription polymerase chain reaction (RT-PCR), which detects viral nucleic acids, is the current gold standard for COVID-19 diagnosis ^7,8^. Although RT-PCR is highly sensitive and specific ^9,10^, it does not detect past infections—RNA is typically only present at high quantities during acute infection—and it does not provide insight into the host’s response to infection ^11^. Serological assays, which detect antibodies induced by SARS-CoV-2, are a crucial supplement to nucleic acid testing for COVID-19 management ^12,13^. Specifically, serological assays are important to track the body’s immune response ^14^, and to potentially inform prognosis ^15^ or immunity status ^12^. Serological assays are also essential for use in epidemiological studies ^16^, and are a critical enabling tool for vaccine development ^17^.

SARS-CoV-2 is an enveloped RNA virus with four structural proteins: spike (S) protein, membrane (M) protein, enveloped (E) protein, and nucleocapsid (N) protein ^18^. As the pandemic unfolded, several serological binding assays were developed including enzyme-linked immunosorbent assays (ELISAs) and lateral flow assays (LFA). These assays measure either the level of total antibody or that of specific antibody isotypes that bind to viral proteins—normally S or N. Several studies have demonstrated promising clinical sensitivity and specificity for ELISA and some LFAs ^19,20^. Furthermore, several ELISAs have been shown to correlate well with neutralizing antibody titers ^21,22^, and thus may be useful clinically and in vaccine development ^23^. However, both ELISA and LFAs have major disadvantages that limit their applicability for COVID-19 management. ELISA requires technical expertise, laboratory infrastructure, and multiple incubation and wash steps, limiting its applicability to settings outside of a centralized laboratory ^24^. On the other hand, LFAs are portable, but they have lower sensitivity and provide qualitative results ^25^, whereas a quantitative readout is preferred for clinical use, research studies, and surveillance applications. Collectively, these shortcomings of ELISAs and LFAs motivate the need for an easily deployable, point-of-care test (POCT) that can be manufactured in large volumes, has quantitative figures of merit equal to laboratory-based tests, and is as easy to use as an LFA.

To address the challenge of creating a user-friendly and widely deployable assay that can detect prior exposure to, and immunological response against SARS-CoV-2, we developed a new multiplexed portable COVID-19 serological assay that is described herein. Our passive microfluidic platform provides sensitive and quantitative detection of antibodies against multiple SARS-CoV-2 viral antigens in 60 minutes with a single test from a single 60 µL drop of blood, plasma, or serum. We chose to quantify the antibody response against three different SARS-CoV-2 antigens because emerging studies have demonstrated that the primary antigenic target of the humoral immune response may inform disease progression and prognosis ^14^. Thus, being able to differentiate the viral targets of antibodies—as we can with our platform—may be especially valuable. Further, our portable test is completely automated and can function independently of a centralized laboratory at the point-of-care. We also show that our test can be easily modified to detect additional protein biomarkers, such as cytokines/chemokines, without compromising the performance of the serological assay, which may provide further clinical insight into disease severity and or patient outcomes ^2,26,27^. Collectively, these attributes suggest that our platform is a valuable tool for COVID-19 management both at the individual patient level (i.e. monitoring patients who may progress to severe disease) and for large-scale epidemiological studies at the population level.

## Results

### The DA-D4 point-of-care test (POCT) for COVID-19 serology

Our strategy to evaluate the antibody response to SARS-CoV-2 is based on the D4 assay platform, developed recently and reported elsewhere ^28^. The D4 platform is a completely self-contained immunoassay platform fabricated upon a “non-fouling” poly(oligoethylene glycol methyl ether methacrylate) (POEGMA) brush, where all reagents needed to complete the assay are inkjet printed directly onto the surface. In previous work, we have used this platform for the detection of several protein biomarkers using a fluorescent sandwich immunoassay format ^28^. Here, we modified the design of the assay to detect antibodies against SARS-CoV-2 using a double-antigen (DA) bridging immunoassay format, which detects total antibody (all isotypes and subclasses). The DA-D4 is fabricated by inkjet printing viral antigens as stable and spatially discrete capture spots. In addition, viral antigens are labeled with a fluorescent tag and are printed nearby on an excipient pad as dissolvable spots. When a sample is added to the assay (**Fig. 1a-i**), the excipient pad dissolves and liberates the fluorescently labeled antigen (**Fig. 1a-ii**), which then diffuses across the polymer brush to the capture spots and labels any antibody that has been captured from solution by the stable capture spots of unlabeled antigen (**Fig. 1a-iii**). The fluorescence intensity of the capture spots is then imaged using a fluorescent detector and scales with antibody concentration in a sample (**Fig. 1a-iv**). Because capture spots of each antigen are printed at spatially discrete locations, this design enables multiplexed quantification of multiple target antibodies using a single fluorescent tag, which greatly simplifies the detector design and assay readout.

**Figure 1.**
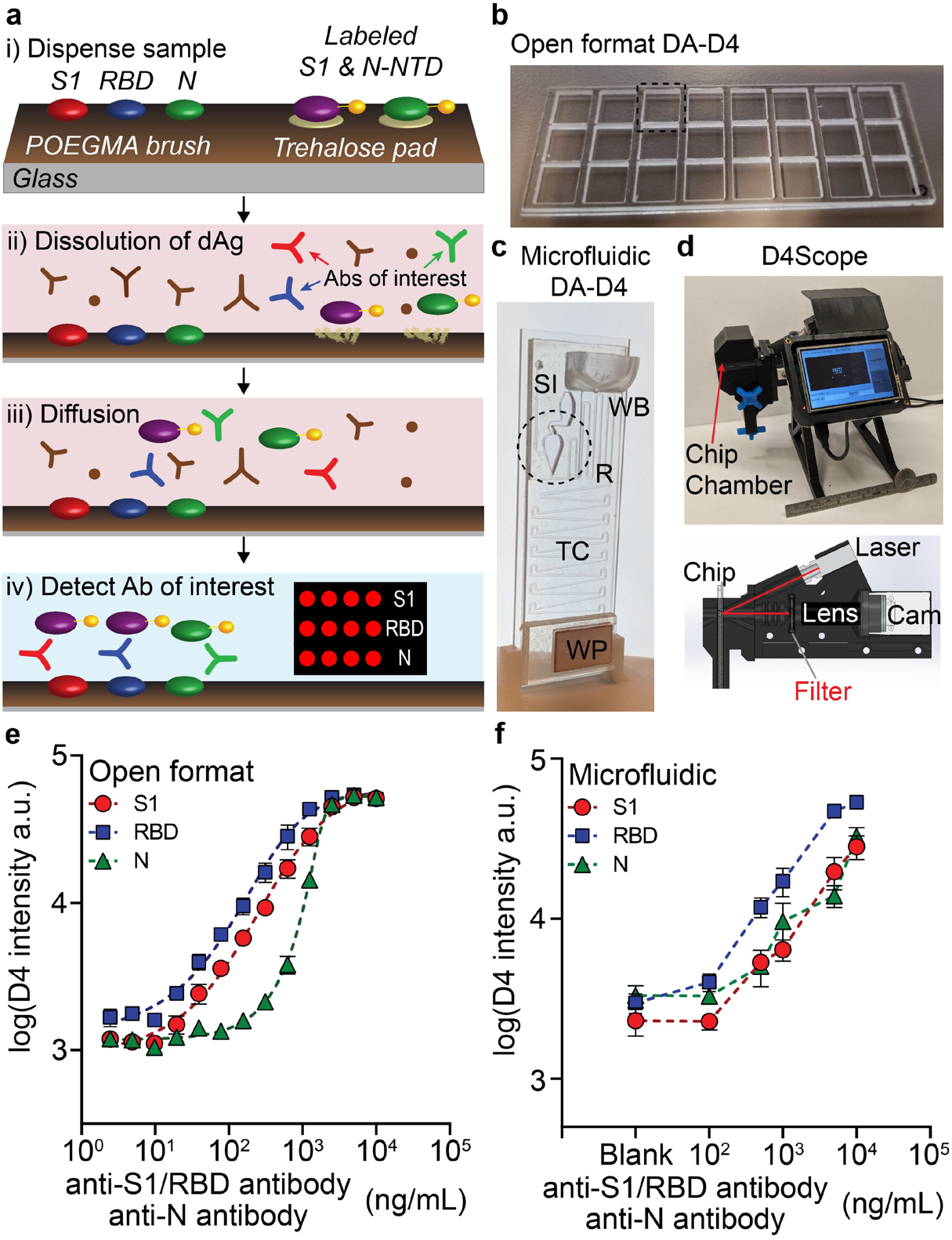
DA-D4 POCT schematic and analytical validation. **(a)** DA-D4 assay chip schematic. S1, RBD, and N capture antigens and fluorescently labeled S1 and N-NTD detection antigens (dAgs) are inkjet printed onto a POEGMA substrate. When a sample is added, dAgs are liberated from the surface due to the dissolution of the underlying trehalose pad. Antibodies targeting each viral antigen then bridge the capture antigens to the dAgs, resulting in a fluorescence signal that scales with antibody concentration. **(b)** Open format DA-D4 with 24-individual assays. **(c)** Microfluidic DA-D4. Sample is added to the sample inlet (SI), filling the reaction chamber (RC) which contains the assay reagents. Wash buffer is added to the wash buffer reservoir (WB) which chases the sample through the microfluidic cassette. The timing channel (TC) sets the incubation time. All liquid is eventually soaked up by the wicking pad (WP) after the incubation process. The size is that of a standard microscope slide. **(d)** D4Scope and cut-away view of the optical path. The microfluidic flow cell is inserted on the left, and pressing a button automates laser excitation, camera exposure, and data output. **(e)** Analytical validation of the open format DA-D4. Antibodies targeting each antigen were spiked into undiluted human serum and incubated for 30 min. Each data point represents the average of three independent runs, and the errors bars represent the standard error of mean (SEM). **(f)** Analytical validation of microfluidic DA-D4. Each data point for an antigen represents the average of four independent microfluidic flow cells and error bars represent the SEM.

To fabricate a serological assay for SARS-CoV-2, nucleocapsid (N), spike S1 domain (S1), and the receptor-binding domain (RBD) of S1 were inkjet printed as the “stable” capture reagents onto POEGMA-coated slides. Our rationale for simultaneously assaying the antibody response towards N, S1, and RBD antigens is that it is not fully understood which epitopes elicit an immune response in all individuals, though they are all believed to be immunogenic ^29,30^ and because studies have shown that the primary target of the immune response may inform disease prognosis ^14^. N is expressed abundantly by SARS-CoV-2 during infection and is highly immunogenic in other coronaviruses ^31,32^. The S protein—composed of the S1 and S2 domains—is exposed on the viral coat of SARS-CoV-2 and plays an essential role in viral attachment, fusion, entry, and transmission ^33^. Because S2 is highly conserved across many coronaviruses and is thus potentially cross-reactive, S1 was chosen for antibody detection ^34^. RBD—the portion of S1 that binds cells expressing viral receptor—is the target for many neutralizing antibodies and is thus a promising antigenic target for serological assays ^34^. **Fig. S1a** shows the layout and dimensions of an open format DA-D4 chip. Each chip contains 24 individual assays with S1, RBD, and N antigens arrayed as separate rows of five identical ∼170 µm diameter spots. Next, fluorescent conjugates of S1—which contains the amino acid sequence for RBD— and the N-terminal domain (NTD) of N (produced in-house, see **Fig. S2** for SDS-PAGE of expression and purification) were mixed 1:1 and inkjet printed as twelve identical 1 mm diameter spots on an identically sized trehalose pad (**Fig S1a**). N-NTD —instead of full-length N— was chosen as the detection reagent because the full N domain can dimerize in solution, potentially leading to a false positive result in the DA format ^35^.

### Analytical validation of the DA-D4 POCT using simulated samples

We first sought to demonstrate that the DA-D4 assay can detect antibodies against recombinantly expressed SARS-CoV-2 antigens. Initially, the analytical performance was characterized using the open format DA-D4 (**Fig. 1b**). This is because the open format DA-D4 assay has been extensively optimized and characterized by our group and has extremely high analytical sensitivity which enables us to determine the figures-of-merit that are theoretically possible for a particular D4 assay. A disadvantage of the open format DA-D4 assay, however, is that it requires a rinse step by the user after incubation of the sample ^28^.

For point-of-care deployment and an improved user experience, we developed —in the course of this study— a new, gravity and capillary driven “passive” microfluidic flow cell that fully automates the assay (**Fig. 1c**). The microfluidic flow cell is fabricated by adhering complementary layers of precision laser-cut acrylic and adhesive sheets onto the functionalized POEGMA substrate (**Fig. S3** and **Fig. S1b** for the print layout). The resulting microfluidic flow cell features a reaction chamber, timing channel, sample inlet, wash buffer reservoir, and wicking pad that automates the sample incubation, sample removal, wash, and drying steps. This simplifies the user experience and limits the possibility of a user incorrectly carrying out the test, as it only requires the user to add the sample and a drop of wash buffer to the cassette. After ∼60 minutes, the cassette is ready for imaging with a custom-built fluorescent detector—the D4Scope (**Fig. 1d**).

The D4Scope is a low-cost (<$1,000), portable fluorescence detector (with dimensions of 7 inches wide, 6 inches tall, 5 inches deep and a weight of ∼5 pounds; see **Fig. S4** for dimensions and image) built from off-the-shelf components and assembled using 3D-printed parts that can image microarray spots with high sensitivity. It utilizes coherent 638 nm red laser light set at an oblique angle (30°) relative to the surface to excite the fluorescently labeled antigens. The fluorescence wavelength emission from the labeled reagents is then sent through a bandpass filter and imaged with a high-efficiency Sony IMX CMOS sensor in a Basler Ace camera (**Fig. 1d**). This setup provides a large field-of-view of 7.4 mm x 5 mm and a fine (raw) lateral resolution of ∼2.4 µm. A user-friendly interface was developed in Python that runs on a 3.5” Raspberry Pi touchscreen to control laser excitation, camera exposure, and image file output (see supplementary information for more details, **Fig. S4**).

To mimic seropositive samples, we spiked commercially available antibodies (with known binding affinity towards SARS-CoV-2 antigens) into undiluted pooled human serum that was collected prior to the COVID-19 outbreak. A dilution series spanning four logs was evaluated on open format DA-D4 chips and yielded a dose-response curve with fluorescence intensities that scaled with antibody concentration and approximated a sigmoidal curve, demonstrating that the assay was responsive to the antibodies of interest (**Fig. 1e**). Within the microfluidic flow cell, the chamber geometry, reagent spacing/alignment, and amount of printed reagent were iteratively optimized to match the performance metrics of the open format DA-D4. Six doses (including a blank) with varying amounts of anti-S1/RBD and anti-N antibodies were prepared and tested in quadruplicate on 24 separate microfluidic flow cells to demonstrate equivalence between the open format (**Fig.1b**) and microfluidic flow cell (**Fig.1c**). In the microfluidic flow cell, the fluorescence intensity of the capture antigens—imaged with the D4Scope—also scaled with antibody concentration, suggesting that the test is responsive to anti-SARS-CoV-2 antibodies (**Fig. 1f**).

### Clinical validation of the DA-D4 POCT

Next, we sought to validate the clinical performance of the DA-D4 POCT in a retrospective study using banked plasma samples from patients with PCR-confirmed COVID-19 who had been admitted to the intensive care unit (ICU) at Duke University Medical Center. A total of 34 COVID-19 positive plasma samples (heat-inactivated) from 19 patients—some of which had longitudinal samples available—and 10 negative samples (collected prior to the COVID-19 pandemic) were tested on the microfluidic DA-D4 and imaged with the D4Scope. The median age of the COVID-19 patients was 55. Of the 19 patients, 10 were female and 9 were male. For most patients, the date of symptom onset was known (29 out of 34 samples), where the average was 20.7 days with a range of 6–48 days. The complete patient profile is provided in **Table S1**.

Antibody reactivity towards all three viral antigens was measured on single microfluidic flow cell for each patient sample. For validation, we conservatively assigned the threshold for a positive test result as three standard deviations above the mean of the negative controls. There was a statistically significant difference between the mean intensity for COVID-19 positive and negative samples (p < 0.0001) for all three markers (**Fig. 2a-c**). 33 out of 34 (97.1%) COVID-19 positive samples tested above the threshold for anti-S1 and anti-N, while 34 out of 34 (100%) tested positive for anti-RBD. All negative controls tested below the threshold for each marker (specificity of 100%). Representative images for a high positive and negative sample are included in **Figure S5**.

**Figure 2.**
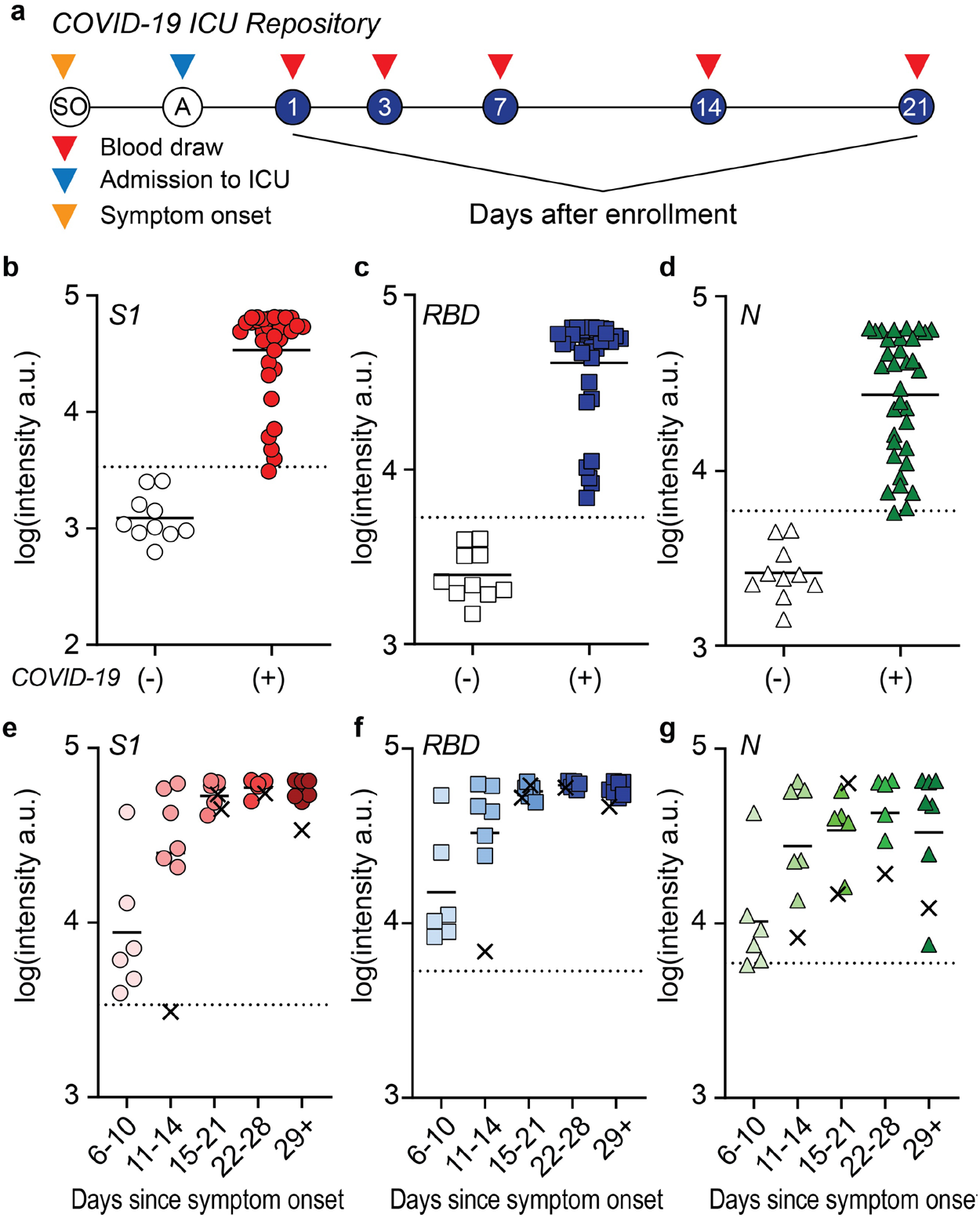
Clinical validation study. **(a)** Study design for COVID-19 ICU Biorepository samples. Patients at Duke University Medical Center were enrolled into the study after admission to the ICU. Blood draws were taken at days 1, 3, 7, 14 and 21 after enrollment until discharge or death occurred. **(b-d)** Aggregated data for 34 positive samples and 10 negative controls tested for antibodies against (b) S1, (c) RBD, and (d) N. Dashed lines represent 3 standard deviations above the mean of the negative controls and solid line represents the mean of each group. **(e-g)** Data from b-d partitioned by days since symptom onset. For 5 samples, date since symptom onset was unknown so days since first positive COVID-19 test was used (marked with an x).

Next, we partitioned the data into five different groups based on days since symptom onset: 6–10 days, 11–14 days, 15–21 days, 22–28 days, and > 29 days (**Fig. 2d-f**). For two patients (five total samples), the date of symptom onset was unknown, hence the day since first positive PCR test result was used instead (these data points are marked with an x). When the data are partitioned by date since symptom onset, the sensitivity for all three markers included in our test increased to 100% for 15 days or longer after symptom onset. For antibodies targeting S1 and RBD, we found a statistically significant difference between the different time groups, as determined by one-way ANOVA (p < 0.0001). Multiple comparisons by Tukey’s post hoc testing revealed that the magnitude of the antibody response to S1 and RBD was higher in all groups after 11 days compared to days 6–10 (p < 0.05) and that there was no statistically significant (p > 0.05) difference between any group after day 11. These results suggest that our assay spans a useful temporal range to detect the dynamic production of antibodies that typically occurs within 2 weeks of symptom onset ^15^. In addition, the patients tested all developed a robust and sustained antibody response against S1 and RBD.

For antibodies targeting N, there was also a statistically significant difference in DA-D4 readout between groups as determined by one-way ANOVA (p < 0.05). However, the production of N-targeting antibodies appears to occur later, as there was no statistically significant difference in the antibody response when comparing days 6 – 10 and 11 – 14 (p > 0.05), but all groups after 15 days were significantly higher than the first time point (p < 0.05). The concentration of N-targeting antibodies also appears to be more variable across all patients, especially at later time points, with some samples testing close to the threshold value. This could be due to the fact that some patients may develop a stronger response against other viral antigens/epitopes (RBD or S1) ^36^ or against an epitope of N not within the NTD, highlighting the importance of testing for antibodies against several antigens simultaneously to maximize test sensitivity and specificity.

We also conducted a proof-of-concept study using whole human blood as the sample source for the microfluidic flow cell, to demonstrate that the DA-D4 assay can be used at the point-of-care or the point-of-sample collection without the need for any sample processing. To do so, we made minor modifications (see section 7 of supplementary information, **Fig.S6**) to the microfluidic timing channel and reaction chamber to account for the non-Newtonian fluid mechanics of whole blood (**Fig. 3a**). Briefly, a gradual slope was added to the reaction chamber to prevent accumulation of red blood cells during washing, and the incubation channel was shortened to account for a reduced flow rate.

**Figure 3.**
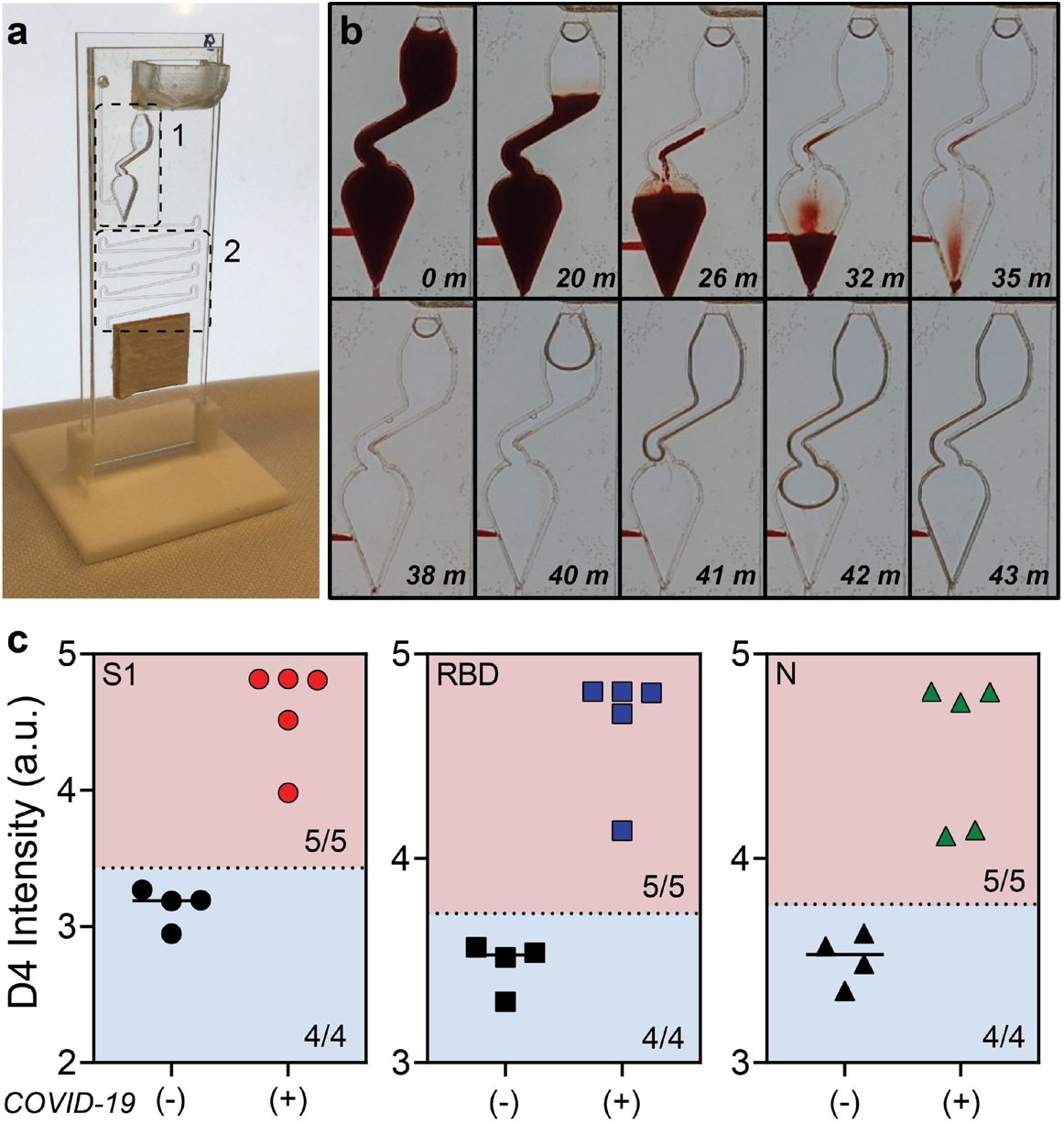
Testing whole blood. **(a)** Modified microfluidic flow cell for testing whole blood. Zone 1: the reaction chamber was modified to prevent red blood cells from collecting in the chamber. Zone 2: The incubation timing channel was shortened to compensate for the slower flow rate of blood and to ensure blood did not clot or clog the channels. **(b)** Time lapse of blood and wash buffer in the reaction chamber. **(c)** Aggregated data for 5 positive samples and 4 negative controls tested for antibodies against S1, RBD, and N. Dashed lines represent 3 standard deviations above the mean of the negative controls. 100% sensitivity (5/5) and 100% specificity (4/4) were achieved for S1, RBD, and N.

Fresh blood was collected in EDTA-coated tubes from four patients with negative COVID-19 antibody status (as determined by ELISA performed by the supplier) and from five patients with confirmed COVID-19 (from new enrollments to the ICU study) (**Table S2**). Each 60 µL blood sample was tested on the microfluidic DA-D4 assay. No complications were observed, such as coagulation of blood that can occur when testing whole blood in microfluidic systems. **Fig. 3b** shows representative images of the reaction chamber, with the time since sample addition noted in the lower right-hand corner of each sub-panel, demonstrating the ability of the microfluidic chip to process whole blood. The antibody response towards S1, RBD, and N from whole blood is shown in **Fig. 3c**. We set the threshold to determine a positive test as three standard deviations above the mean of the negatives. All negative samples tested as negative, and all positives tested above the threshold. These preliminary results suggest that the microfluidic DA-D4 assay is capable of detecting anti-SARS-CoV-2 antibodies in whole blood, so that the assay can be carried out immediately at the point of sample collection without the need for transport to a centralized laboratory for sample processing into serum or plasma and subsequent testing.

### Monitoring antibody levels longitudinally

Having demonstrated the high clinical sensitivity and specificity of the microfluidic DA-D4 assay for detection of antibodies against SARS-CoV-2 antigens, as well as the ability to detect changes in antibody levels with time, we next sought to track individual patients to track their seroconversion. To accomplish this, we tested longitudinal plasma samples from six individual patients (**Fig. 4a-f**). Across all six patients, the antibody response was initially low for the first time point tested and then increased and plateaued at later time points, consistent with the antibody dynamics reported in other studies ^15,37,38^. The DA-D4 readout for antibodies targeting S1 and RBD appeared to saturate by the second time point—typically 2-3 weeks post symptom onset—suggesting that each patient mounted a strong and robust immune response that was sustained over time. For N, the dynamics were slower in one patient (#1) and did not fully saturate in another (#3), providing insight into the primary target of the antibody response in those patients. In general, patients with severe COVID-19 often develop very high antibody titers ^37^, which is reflected in this ICU patient sample set by saturated signals at later time points. However, we were still able to measure seroconversion and antibody kinetics in each patient, suggesting that the DA-D4 is a useful tool for monitoring the immune response. The earliest time points for each patient were also still elevated relative to the negative controls, indicating that we may have been able to detect seroconversion earlier, had samples from earlier time points been available. For patients later in disease progression with high antibody titers, dilutions could be performed to adjust the concentration into the linear range of the assay. Testing a sample at various dilutions would also allow us to calculate specific antibody titers, which we are not able to do from a single undiluted sample.

**Figure 4.**
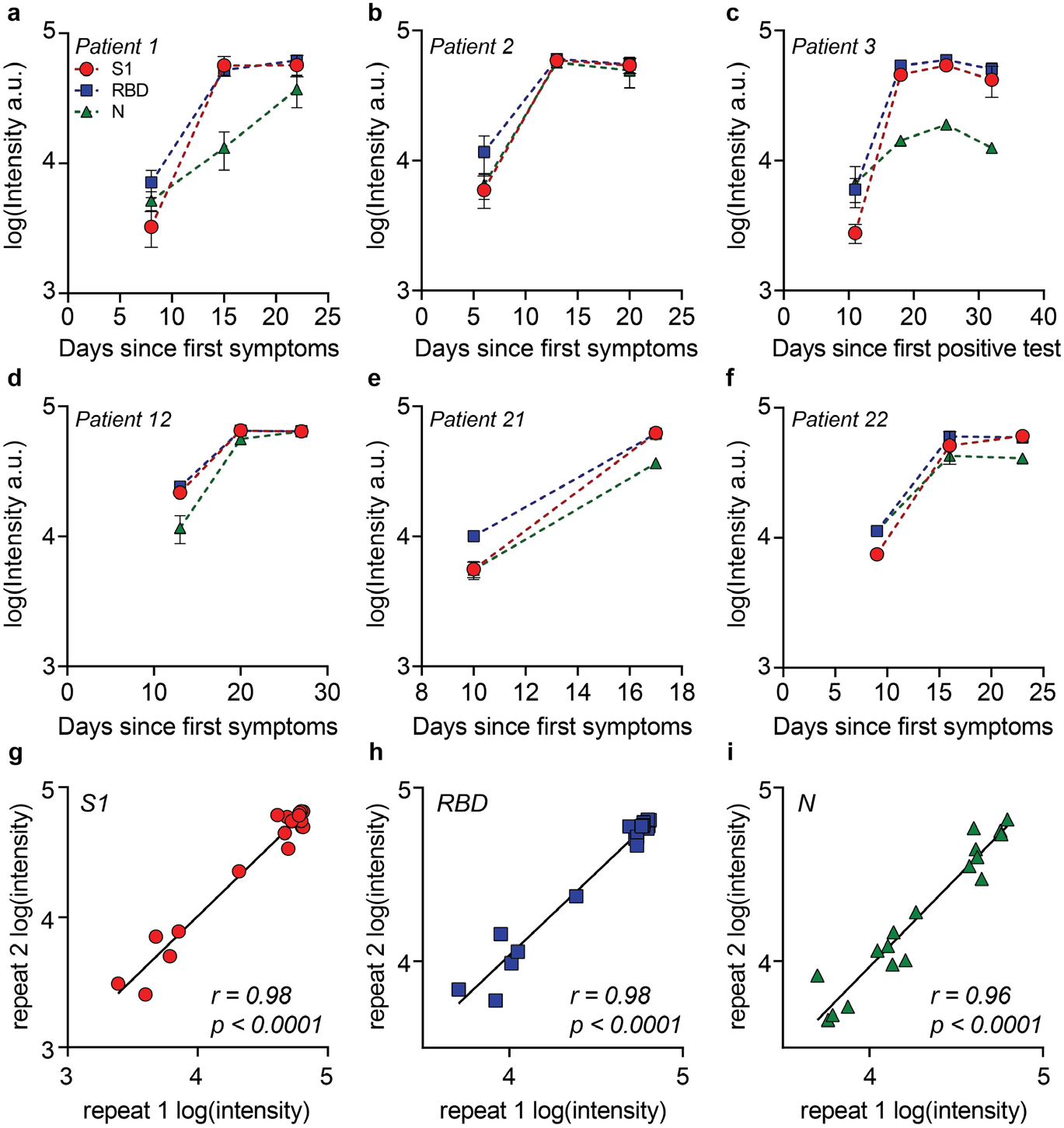
Longitudinal antibody tracking. **(a-f)** Six patients were tracked across multiple time points for antibodies targeting S1, RBD, and N. For patient 3 (c), date since symptom onset was unknown so days since first positive test was used. Each data point represents the average of two independent chips (with SD) run by separate users. **(g-i)** Data from parts a-f for each repeat. There is a strong correlation between each repeat for (g) S1, (h) RBD, and (i) N, with a Pearson r of 0.98, 0.98 and 0.96, respectively (p < 0.0001).

Each sample in the longitudinal study was tested in duplicate on different days and by a different user to characterize the reproducibility and robustness of our platform (**Fig. 4g-i**). We found a strong correlation for each marker, with a Pearson’s r correlation of 0.98, 0.98, and 0.96 for S1, RBD, and N, respectively. The high correlation between replicates further emphasizes the quantitative nature and reproducibility of our platform for profiling the immune response to SARS-CoV-2.

### Concordance with neutralizing antibody titers

We next compared the performance of the DA-D4 with a microneutralization assay that monitors functional neutralization of SARS-CoV-2 via neutralizing antibodies binding to the RBD. All six patients that we tracked longitudinally developed robust neutralizing antibodies, and the microneutralization titer was strongly concordant with DA-D4 assay readout for antibodies targeting S1 and the RBD of S1 (**Fig. 5a-f**). Furthermore, a concordance analysis of the DA-D4 assay with the microneutralization assay for antibodies targeting S1 and RBD showed a strong correlation across all plasma samples tested (**Fig. S7a, b**), as determined by a Pearson r > 0.70 (p < 0.0001). For antibodies targeting N, the concordance between the two assays was not as strong, with only a moderate correlation between the DA-D4 results and microneutralization data (**Fig. S7c**). This is expected, as N resides inside the capsid of SARS-CoV-2 and is not relevant for functional neutralization ^39^. This is also reflected in the longitudinal sample set. For example, patient #1 at day 15 after symptom onset has strong neutralizing antibodies, as seen by the microneutralization assay, despite a weak overall antibody level for N. Although future studies are required to validate the ability of neutralizing antibodies to confer protection, these results suggest that the DA-D4 assay could be used as a supplement to live virus neutralization assays, which typically require >48 hours and biosafety level 3 containment.

**Figure 5.**
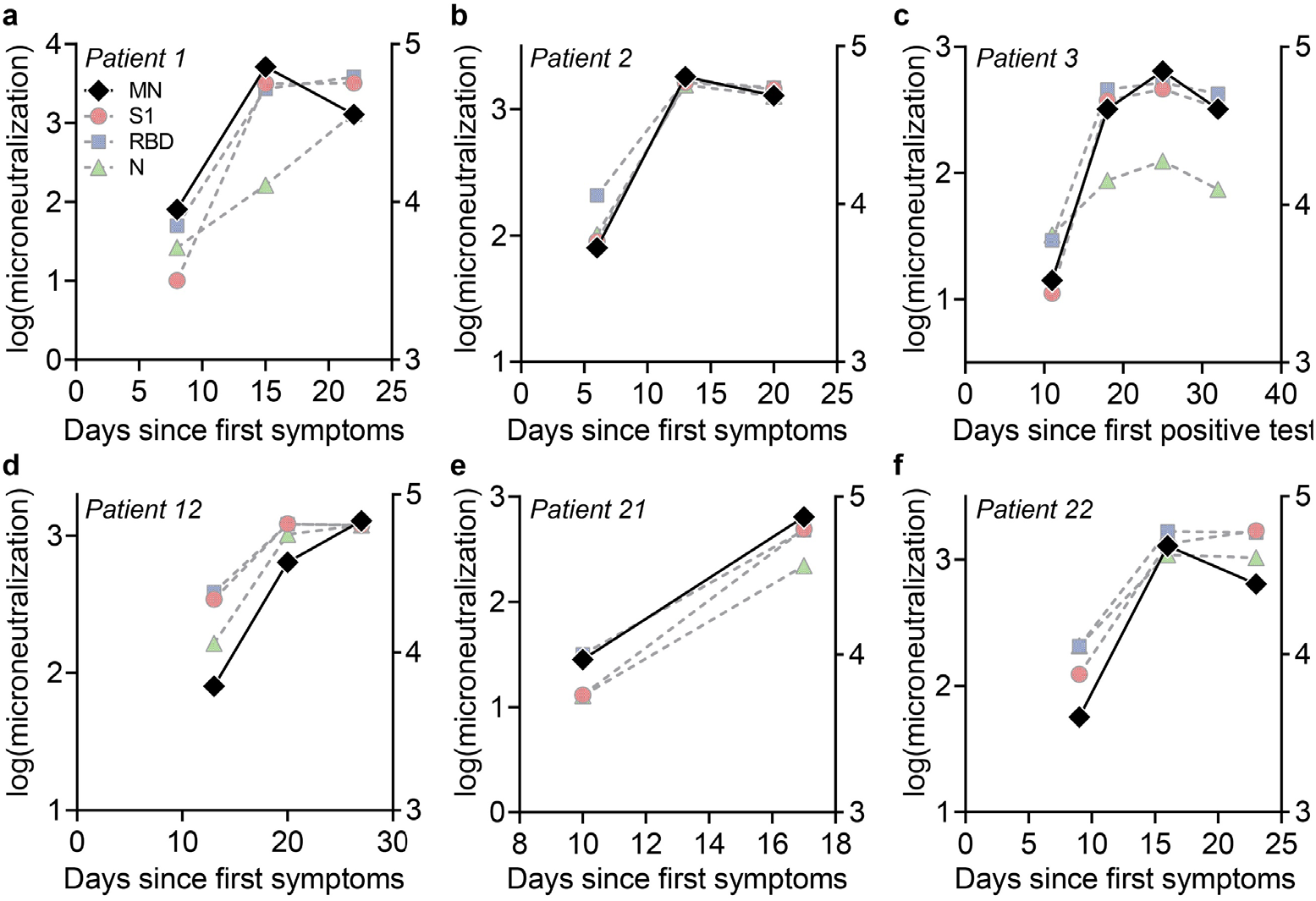
Correlation to microneutralization assay. **(a-f)** Microneutralization assays were performed on each longitudinal sample (black diamonds). Microneutralization titer is plotted on the left axis superimposed against the antibody data from figure 3 (plotted on the right axis).

### Profiling prognostic biomarkers concurrent with serological testing

Finally, we investigated the feasibility of detecting a prognostic protein biomarker concurrent with serological profiling. This is motivated by the fact that others have identified potentially prognostic biomarkers that correlate well with disease severity and patient outcomes ^40,41^. Therefore, tracking antibody levels alongside prognostic biomarkers may provide clinically relevant information to inform interventions in the ICU for patients with a high probability of a poor outcome. As proof-of-concept, detection of IFN-γ–induced protein 10 (IP-10, CXCL10)—a chemokine that recruits inflammatory cells to the site of inflammation and which has been shown to be elevated in severe disease and correlates with patient prognosis ^27,41^—was integrated into the DA-D4 assay using a traditional sandwich immunoassay approach, as described previously ^28^.

Prior to testing patient samples, we sought to confirm that the multiplexed serological assay is compatible (not cross-reactive) with the IP-10 sandwich assay. To do so, we fabricated open format chips containing all necessary reagents for both COVID-19 serology and human IP-10 detection. First, we prepared a 15-point dilution series of recombinant human IP-10 spiked into fetal bovine serum (FBS)—spanning the relevant physiological range for COVID-19 patients identified elsewhere ^41^—and added samples to chips in triplicate in the absence of antibodies targeting SARS-CoV-2 antigens. We observed a dose-dependent behavior for IP-10 response with a low limit-of-detection of 0.12 ng/mL ^42^ and minimal reactivity for SARS-CoV-2 capture antigens, confirming that the IP-10 assay components do not cross react with the serology components (**Fig. 6a**). Next, we prepared a dilution series of simulated seropositive samples and added them to the open format chips. Across all concentrations of anti-SARS-CoV-2 antibodies, IP-10 capture antibody intensity was close to baseline, thus confirming that the serology components do not interfere with the IP-10 detection assay (**Fig. 6b**).

**Figure 6.**
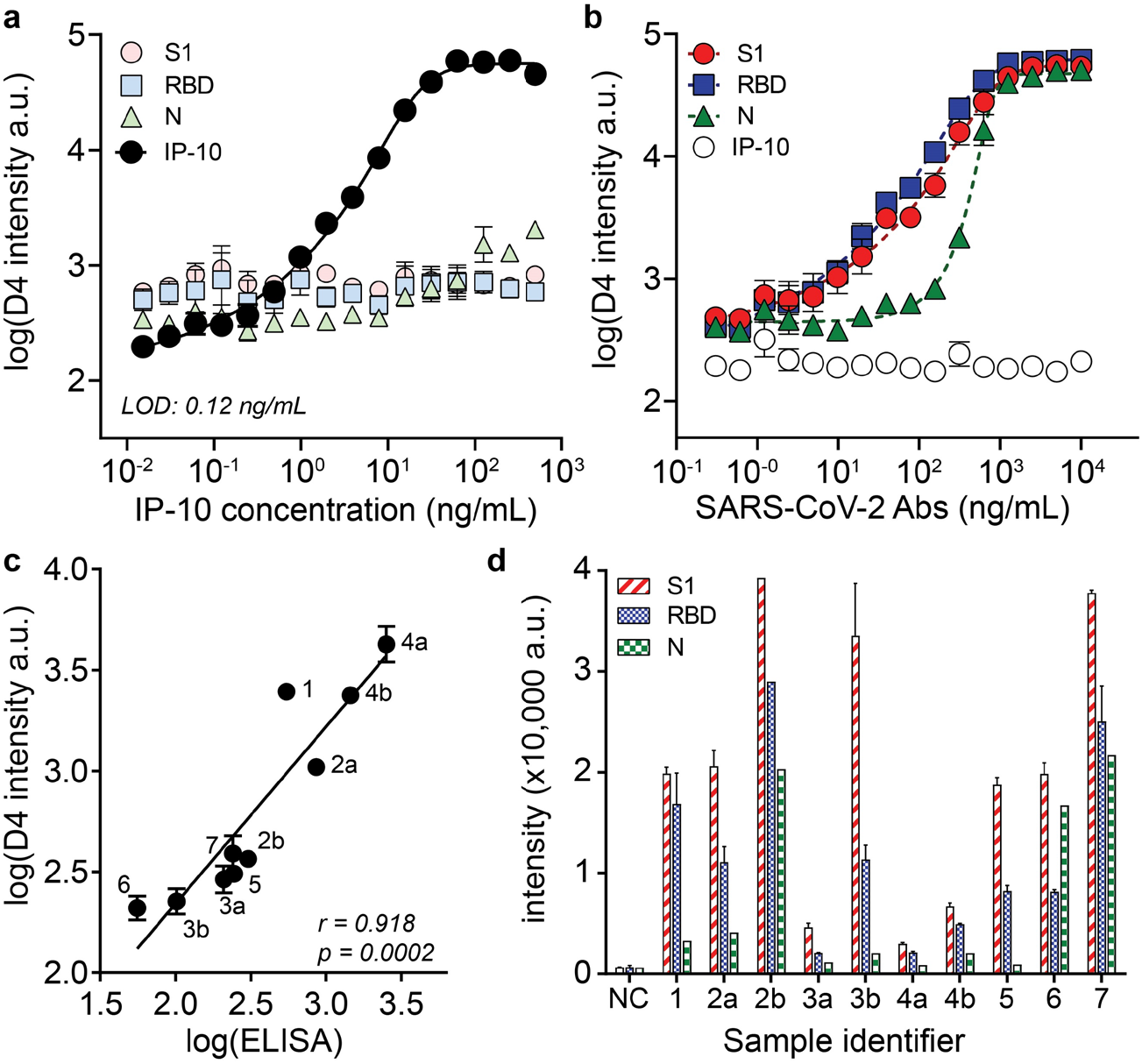
Combined prognostic biomarker and serology detection. **(a)** Dose-response curve for recombinant IP-10 spiked into FBS. Each data point represents the average (n = 3) and error bars represent the SEM. The limit-of-detection (LOD) for IP-10 is 0.12 ng/mL. **(b)** Dose-response curve for anti-SARS-CoV-2 antibodies spiked into FBS. The highest concentration is 10 µg/mL of anti-S1/RBD and 10 µg/mL of anti-N antibodies. **(c)** Correlation between DA-D4 readout for IP-10 with an ELISA assay performed separately. Samples with a letter designate samples from one individual at different time points, where b occurs later in disease than a. All samples were tested in duplicate on the DA-D4 (with SD shown) except 2b (due to insufficient volume). **(d)** Antibody reactivity against S1, RBD, and N for sample tested in part c (with SD shown). NC = negative control pooled healthy plasma.

Having confirmed the compatibility of the IP-10 assay with multiplexed serology in the open D4 format, we next sought to test the performance of our assay in patient samples. Ten COVID-19 positive plasma samples (from 7 patients) were procured from the ICU biorepository and were added undiluted to open format chips and then quantitatively assessed by the DA-D4. Separately, serum samples from the same patients were evaluated in parallel via LEGENDplex™ ELISA assay kits which report IP-10 concentration in pg/mL. We observed a strong positive correlation between the DA-D4 assay for IP-10 with ELISA across all 10 pairs of measurements, with a Pearson’s r of 0.918 (p = 0.0002, 95% CI: 0.68 to 0.98) (**Fig. 6c**). We also tested for antibody reactivity towards S1, RBD, and N from the same samples and an additional sample of healthy pooled plasma (pre COVID-19 negative control) (**Fig. 6d**). Although we did not observe a strong relationship between antibody and IP-10 levels (data not shown), we did observe that in the patients for which we tested multiple samples, IP-10 decreased over time, while the levels of antibodies increased.

Overall, these results clearly show that the D4 assay format can simultaneously detect antibody response to foreign native SARS-CoV-2 antigens and a native protein biomarker from undiluted patient plasma. One of the benefits of detecting anti-SARS-CoV-2 antibodies from undiluted samples is that the sensitivity of the protein detection assay is not reduced because of dilution, allowing us to detect chemokines and cytokines—which are present at very low concentrations even during disease state—directly from complex biological milieu. Detection of additional prognostic biomarkers could also be implemented on the same chip, as long as there is no cross-reactivity between the assay reagents for serology and prognosis. Interestingly, a recent study found that the ratio of IL-6 to IL-10 can be used to guide clinical decision making ^43^, which we plan to measure in the next generation of this assay.

## Discussion

As the COVID-19 pandemic unfolded, countries around the globe grappled with developing streamlined systems for diagnosis of acute infection using nucleic acid detection methods. Although there remains an urgent need for rapid and sensitive point-of-care tests for acute diagnosis, developing accurate and reliable serological assays has been deemed an equally important endeavor to complement existing diagnostic strategies ^12,44^. The challenge with developing an easy-to-use serology assay that can be broadly disseminated, but that performs as well as centralized laboratory-based methods is highlighted by the large number of ELISA and LFA tests that have been developed. While LFAs are portable and easy-to-use and ELISAs are quantitative and highly sensitive, there remains a need for a technology that can merge the best attributes of each format.

The DA-D4 POCT is a promising platform to supplement existing diagnostic technologies to manage the COVID-19 pandemic because it marries the best attributes of LFAs and ELISAs— it is quantitative, easy to use, widely deployable, requires only a single 60 µL drop of blood, and can be performed with minimal user intervention. The SARS-CoV-2 DA-D4 assay can be used to measure antibody kinetics and seroconversion at the individual patient level directly from unprocessed blood or plasma. This test is highly sensitive and specific and is potentially suited for epidemiological surveillance at the population-level using low cost microfluidic cassettes that can be transported and stored for an extended period of time without a cold chain, and that require minimal user intervention to carry out the assay, which provide a quantitative readout using a low cost, hand-held detector.

We show a strong correlation between the DA-D4 assay readout (for S1 and the RBD of S1) and neutralizing antibody titers, suggesting that this test may be useful in understanding efficacy and durability of natural or vaccine-induced humoral immunity, and to potentially inform disease prognosis and population-level immunity. We also demonstrate that an additional prognostic biomarker can be easily incorporated into the test, which may be useful for monitoring disease severity and predict clinical outcomes. Combined, these attributes suggest this platform may also be useful on the individual patient level to aid in clinical decision making. While the results presented here mainly highlight the performance of the microfluidic chip, the open format architecture with up to 24 individual assays per glass slide may be useful for scenarios where higher throughput testing is demanded. The open format still has advantages compared to traditional ELISA because the open format only requires a single incubation step and one wash step, which reduces the hands-on time and equipment complexity required to complete the assay.

The DA-D4 has additional features that synergize to deliver a highly desirable serological assay. First, the double-antigen sandwich format (i.e. antibody bridging) has advantages over other serological assay formats. Because total antibody is detected rather than a single antibody isotype or subclass, seroconversion in patients can be detected earlier, which reduces the chances of a false negative result due to a test being administered too early in disease ^38^. Furthermore, because the labeled reagent does not have species specificity, the single assay kit could be used in pre-clinical vaccine development studies to measure antibody responses in experimental animals ^23^. The lack of species-specific detection antibodies also reduces the risk of high background signal caused by non-specific antibodies binding to the surface and subsequently being labeled ^45^.

Second, all reagents needed to complete the assay are incorporated onto the non-fouling POEGMA brush, which eliminates virtually all non-specific protein adsorption and cellular adhesion, thereby enabling an extremely low LOD directly from undiluted samples ^46-48^. Although many serological assays often dilute samples, the ability to test undiluted samples is advantageous, especially when combined with prognostic biomarker testing where dilution of low concentration analytes can lead to an undetectable signal. Testing multiple dilutions can still be performed using our test when antibody levels become high, which could be used to calculate specific titers. POEGMA also acts as a stabilizing substrate for printed reagents, enabling long term storage of chips without a cold chain ^28^. In this study, results were generated over the course of three months from the same batch of tests stored in silica desiccated pouches at room temperature and ambient humidity.

Third, this platform can be easily multiplexed, which can be used to capture a more detailed picture of the host immune response to SARS-CoV-2 infection by quantifying the antibody level induced to multiple viral antigens —in this case N, S1, and S1-RBD– from a single sample without sacrificing ease-of-use. This is because each viral antigen is deposited at a spatially discrete location, which allows for a single fluorescent tag to be used during fluorescence imaging of the chip, thereby simplifying assay readout compared to other multiplexing technologies such as Simoa or Luminex assays which rely on multiple different reporter molecules and a more complex readout ^14,49^. This method also allows us to simultaneously measure the concentration of potential prognostic biomarkers directly from plasma ^26,27^ without compromising the performance of the multiplexed serological assay. To the best of our knowledge, there are currently no tests on the market that can probe for antibodies against multiple viral antigens and prognostic protein biomarkers simultaneously.

Fourth, this platform is designed for point-of-care deployment because it requires a single drop of blood that is readily obtained from a fingerstick. This droplet is injected into the sample port of a gravity driven microfluidic chip that requires no further user intervention beyond the concurrent addition of a few drops of wash buffer into a separate port. The assay runs by itself under the action of gravity and capillary action until all the fluid is drained from the microfluidic path by the absorbent pad at the bottom of the cassette, which fully absorbs and contains all liquid. The microfluidic chip relies only upon capillary action and gravity to drive fluid flow, which eliminates the need for pumps, valves, or actuators, and reduces the complexity and cost of the assay. This enables the assay to be read out at the point of sample collection using the D4Scope—a highly sensitive and inexpensive handheld detector developed to work with the microfluidic chip. The D4Scope images a chip and provides a quantitative readout in less than 5 seconds, does not require an external power source or laboratory infrastructure, and can wirelessly transmit the results to a remote server over Wi-Fi. While smartphone-based diagnostics are becoming more popular, a benefit of this platform is that it does not rely on smartphone hardware and software, which change rapidly. Combined, these attributes make our platform ideal for providing ELISA-like sensitivity and quantitation with the ease-of-use and scalability of LFAs.

Where might this point of care assay for COVID-19 serology and prognosis be useful? Serial quantification of antibody response and prognostic biomarkers would be most useful to monitor symptomatic and severe cases where use of available therapeutics, such as antiviral or monoclonal therapies, is indicated. Further, it could be used to screen for patients with poor antibody responses who may benefit from convalescent plasma or monoclonal antibody therapy. We believe that this platform has potential utility in point-of-care settings such as ICUs, urgent care clinics, and at the point-of-use—at locations where periodic surveillance of healthcare workers and other essential workers in close proximity to others for extended periods of time such as assembly-line manufacturing or food processing plants— is desirable to assist in tracking clusters of disease and epidemiological studies. This platform could also be used as an inexpensive tool to study the longitudinal dynamics of antibody levels to inform re-infection potential, as coronavirus immunity often lasts only ∼6 months ^50^. Similarly, it could be used to monitor vaccine-induced humoral immunity, which could help determine if boosters are needed in certain vaccinated individuals. This technology is suitable for low-resource settings across the globe, where eliminating the need for sample storage and transport to a centralized testing facility, and the attendant cold chain, is desirable, and where access to expensive, high-throughput clinical analyzers that process large volumes of serology and other sandwich immunoassays is limited. Similarly, remote and austere settings —such as the field-forward position of the military or other remote locations where pandemics often emerge— can also benefit from this platform, as the testing is carried out with a disposable cassette and a low-cost, light-weight, and handheld detector whose production can easily be scaled up to enable wide-spread and dispersed deployment.

While the results presented here are promising, there are several issues identified during this study that require further investigation prior to its deployment. First, our cohort of individuals with SARS-CoV-2 infection consisted of adults with clinically significant disease, which is not representative of the entire spectrum of COVID-19 disease severity. These samples were chosen to demonstrate proof-of-concept of the DA-D4 assay and because these samples were locally available through an existing biobank. We recognize that a larger sample size that spans the disease severity spectrum is required to develop a more robust measure of sensitivity and specificity of the DA-D4 serology test for SARS-CoV-2. Similarly, we were not able to match demographics in our negative control group, which may have introduced confounding variables in our analyses. Furthermore, several of the samples we tested saturated the readout of our assay, which limits the dynamics we can measure once high antibody titers are achieved. This limitation could be addressed by testing individual samples on separate microfluidic chips at various dilutions, which would effectively increase the dynamic range of our assay and yield more precise quantitative titer. Additionally, because of the double antigen design of our assay, we are also not able to discriminate between specific antibody subclasses or isotypes, which has been shown to be important for other diseases. Despite these limitations, we believe our assay is well poised to complement existing diagnostic solutions once additional validation studies encompassing larger patient cohorts are completed.

In summary, we have developed a COVID-19 serological assay that merges the benefits of LFAs and ELISAs. We used this test to simultaneously measure the antibody levels for multiple viral antigens and a potential prognostic biomarker directly from plasma and whole blood. For COVID-19 management, our platform may be useful to better understand patient antibody responses, provide actionable intelligence to physicians to guide interventions for hospitalized patients at the point-of-care, to assess vaccine efficacy, and to perform epidemiological studies. Further, our platform is broadly applicable to other diseases where sensitive and quantitative antibody and or protein detection is desirable in settings without access to a centralized laboratory. Overall, we believe that our platform is a promising approach to democratize access to laboratory quality tests, by enabling rapid and decentralized testing with minimal user intervention to locations outside the hospital.

## Materials and methods

### DA-D4 assay

The DA-D4 assay is based on the design of the D4 immunoassay, reported elsewhere ^28^. Briefly, a polymer brush composed of poly(oligoethylene glycol methyl ether methacrylate) (POEGMA) was “grafted from” a glass slide by surface-initiated atom transfer radical polymerization ^47^. Recombinant SARS-CoV-2 proteins were then printed onto POEGMA-coated slides as capture and detection spots. Capture spots of the following proteins were printed as ∼170 µm diameter spots using a Scienion S11 sciFLEXARRAYER (Scienion AG) inkjet printer: Spike S1 (Sino Biological, cat# 40591-V05H1), Spike RBD (Sino Biological, cat# 40592-V02H), and Nucleocapsid protein (Leinco, cat# S854). Each protein was printed as a row/column of five identical spots. Next, 12 excipient pads of trehalose with 1.6 mm spacing were printed from a 10% (w/v) trehalose solution in deionized water around the periphery of the capture antigen array using a BioDot AD1520 printer (BioDot, Inc.). To print the detection reagents, S1 (Sino Biological, cat# 40591-V08H) and N-NTD (produced in-house), were first conjugated to Alexa Fluor 647 (per the manufacturer’s instructions) and then detection spots, of the fluorescent protein conjugates of these proteins were printed on top of the excipient pads as twelve 1 mm diameter spots. A schematic of the chip that shows the spatial address and dimensions of the capture spots, trehalose pad and detection spots is sown in **Figure S1**. After printing and final assembly, D4 chips were stored with desiccant until use. The amount of reagent deposited for the open-format and microfluidic format was identical, with the only difference being the relative spot placement (**Fig. S1a, b**). For DA-D4 assays that also detected IP-10, an additional column of five spots of capture antibody (R&D systems, cat# MAB266) was included and anti-IP-10 detection antibody (R&D systems, cat# AF-266) was included in the detection cocktail for the open format chips.

### Fabrication and analytical testing of open format DA-D4

Open format slides were prepared by adhering acrylic wells to each slide, which splits one slide into 24 independent arrays (see **Fig. S1a** for a schematic and **Fig. 1b** for an image). To validate the analytical performance of the test, dose-response curves were generated using antibodies targeting SARS-CoV-2 antigens (Sino Biological, cat#: 40143-MM05, 40150-D001, and 40150-D004) spiked into undiluted pooled human serum. Open format chips were incubated with a 15-point dilution series (run in triplicate) for 30 minutes, briefly rinsed in a 0.1% Tween-20/PBS wash buffer and then dried. Arrays were imaged on an Axon Genepix 4400 tabletop scanner (Molecular Devices, LLC).

### Fabrication and analytical testing of microfluidic DA-D4

The microfluidic chip fabrication process is described in detail in the supplementary information section 3. Briefly, the microfluidic chip was fabricated by adhering complementary layers of precision laser-cut acrylic and adhesive sheets onto the POEGMA substrate that had been functionalized with the relevant capture and detection reagents. The resulting assembly features a reaction chamber, timing channel, sample inlet, wash buffer reservoir, and wicking pad that automates the sample incubation, sample removal, wash, and drying steps. Simulated doses were prepared using antibodies targeting SARS-CoV-2 antigens (Sino Biological, cat#: 40143-MM05, 40150-D001, and 40150-D004) spiked into undiluted pooled human serum. Six doses (including a blank) were tested on the microfluidic DA-D4 in the following way: (1) The user dispenses 60 µL of sample into the sample inlet using a pipette. (2) The user dispenses 135 µL of wash buffer into the wash reservoir of the cassette using a pipette. (3) The user waits 60 minutes for the cassette to run to completion. During this time, (a) fluorescently labeled antigens dissolve and form sandwiches with the antibodies of interest and the immobilized capture antigen in the reaction chamber. (b) A small volume of sample traverses the timing channel, which governs the incubation time. (c) The sample reaches an absorbent pad situated at the end of the timing channel that rapidly wicks away all sample from the reaction chamber, ending incubation. (d) As the sample clears, wash buffer enters the reaction chamber removing residual sample and unbound reagent before it is also wicked away leaving a cleaned and dry imaging surface. We observed less than a +/- 10% variation in the designed 23-minute incubation time for the data presented in Figure 1f. The remaining difference in time accounts for washing and drying time. (4) The cassette is ready for analysis on the D4Scope. The vertical orientation of the cassette works in conjunction with the POEGMA brush to maintain low background fluorescence. Cellular and other sample debris can collect on the brush surface due to gravitational forces, even if no binding is occurring. The vertical orientation ensures that these debris fall harmlessly towards the timing channel during the wash step. This proved especially important when testing with undiluted human whole blood samples.

### Clinical testing of microfluidic DA-D4

The same testing procedure as described in “*Analytical testing of microfluidic DA-D4*” was used to test both the plasma and whole blood clinical samples. For both cases, sample was added directly to the microfluidic DA-D4. For blood, a variation in the microfluidic design was used and is described in supplementary information section 7. This required the use of 200 µL of wash buffer. All other procedures remained the same.

### D4Scope fabrication and operation

The D4Scope design, fabrication, and assembly is described in detail in the supplementary information section 5. Briefly, the D4Scope’s optical elements – the laser, bandpass filter, lens, and camera—and processing elements – the Raspberry Pi 4, touchscreen, and cabling—are mounted in a custom 3D printed chassis. Fully assembled, it weighs ∼5 pounds. The D4Scope can be powered either through a portable battery pack or wall power. Once connected to the power source, the D4Scope automatically runs our custom imaging Python program. The user removes the light protection cover from the cassette loading port and slides the microfluidic cassette with glass side towards the detector. The light protection cover is then replaced enclosing the cassette. The user is then prompted to enter the sample ID # and chip ID # using either the touchscreen or optional attached keyboard and mouse.

The D4Scope has two fine adjustment knobs on the cassette loading port that allow for precise vertically and horizontally movement of the cassette relative to the laser source to ensure that the DA-D4 array is perfectly centered with the excitation source. Each array has co-printed two control spots that will always be uniformly bright across all tested samples and align with two super-imposed alignment cross hairs on the live video-feed of the D4Scope. Using the “Toggle video” function on the UI activates the laser and camera to provide a live view of the imaging area for this alignment. Once aligned, the “Toggle video” function can be pressed again to end the live view, and the “Capture image” function can be used to collect and save the resulting image onto the on-board hard-drive and optionally to a cloud-based server defined by the end-user. The live-view feature should be used sparingly to prevent photo-bleaching of the sample. For this study we manually analyzed the resulting fluorescence intensity using Genepix Analysis software. However, we have developed an algorithm for automatic analysis of spot intensity and instantaneous results on our open-format platform, which will be reported elsewhere.

### Patient samples

De-identified heat-inactivated EDTA plasma samples (57°C for 30 minutes) were accessed from the Duke COVID-19 ICU biorepository (Pro00101196, PI Bryan Kraft) via an exempted protocol approved by the Duke University Institutional review board (Pro00105331, PI Ashutosh Chilkoti). Briefly, eligible patients included in the repository were men and women ages 18 years and above that were admitted to an adult ICU at Duke University Hospital with SARS-CoV-2 infection confirmed by PCR testing. Samples were collected on study days 1, 3, 7, 14, and 21. In addition to biological samples, clinical data on these patients were also collected including demographics, laboratory data, and clinical course. All data reported in this paper were obtained with patient samples from the Duke ICU Biorepository and this study was performed in collaboration with Biorepository team.

Negative control plasma samples were collected under a normal blood donor protocol (Pro00009459, PI Tony Moody) and range in collection dates from 2014 to 2019 (prior to the COVID-19 outbreak). All patient information, including demographics, is unknown to the investigator team. These samples were accessed via an exempted protocol approved by the Duke University Institutional review board (Pro00105331, PI Ashutosh Chilkoti).

Blood was either purchased commercially (Innovative Research, Inc.) or accessed from the ICU biorepository (Pro00101196, PI Bryan Kraft) in EDTA-collection tubes and was tested within 48 hours of sample collection.

### Live SARS-CoV-2 Microneutralization assay (MN)

The SARS-CoV-2 virus (Isolate USA-WA1/2020, NR-52281) was deposited by the Centers for Disease Control and Prevention and obtained through BEI Resources, NIAID, NIH. SARS-CoV-2 Micro-neutralization (MN) assays were adapted from a previous study ^51^. In short, plasma samples are diluted two-fold and incubated with 100 TCID50 virus for 1 h. These dilutions are transferred to a 96 well plate containing 2×10^4^ Vero E6 cells per well. Following a 96 h incubation, cells were fixed with 10% formalin and CPE was determined after staining with 0.1% crystal violet. Each batch of MN includes a known neutralizing control antibody (Clone D001; SINO, CAT# 40150-D001). Data are reported as the inverse of the last dilution of plasma that protected from CPE, log10 transformed.

### IP-10 experiments

Open format DA-D4 slides were fabricated as described above using all reagents needed for antibody detection and IP-10 detection. Citrated plasma samples from 10 patients were procured from the ICU biorepository. 60 µL of each sample was added to two separate DA-D4 chips, incubated for 30 min, and the chips were then rinsed using 0.1% Tween in 1x PBS. All slides were scanned with the Genepix tabletop scanner.

IP-10 levels were measured using the LEGENDplex™ Human Proinflammatory Chemokine Panel (13-plex) and LEGENDplex™ Human Anti-Virus Response Panel (13-plex) obtained from BioLegend. Assays were performed with patient serum per the manufacturer’s instructions. The assay was performed using a Beckman Coulter CytoFLEX flow cytometer and data processing was performed using BioLegend’s Bio-Bits cloud-based software platform. Each sample was tested in triplicate, and the results are reported as mean of these triplicates.

### Statistical analysis

Statistical analysis was performed using GraphPad Prism version 8.4.1 (GraphPad Software, Inc). All data were log transformed for analysis. To establish statistical significance between two groups (Fig. 2a-c), unpaired t-tests were used. When comparing multiple groups, a one-way ANOVA followed by Tukey’s post-hoc multiple comparisons test was used. Pearson r correlation was used to assess the degree of correlation between measurements.

## Data Availability

Data are included in the manuscript and supplementary information

## Acknowledgments

AC acknowledges the support of the National Science Foundation (Grant No. CBET202936);the National Cancer Institute through grants P30-CA014236, R01-CA248491, UH3-CA211232; Department of Defense United States Special Operations Command (Grant No.W81XWH-16-C-0219); Defence Academy of the United Kingdom (Grant No. ACC6010469); and the Combat Casualty Care Research Program (JPC-6) (Grant No. W81XWH-17-2-0045). BDK receives funding from NHLBI (K08HL130557). We thank Dr. David Montefiori for providing laboratory space to complete the clinical validation studies. We also thank Rebecca Sahm for completing live SARS-CoV-2 microneutralization assays, which were performed in the Virology Unit of the Duke Regional Biocontainment Laboratory, which received partial support for construction from the NIH/NIAD (UC6AI058607; GDS). We thank the nurses in the intensive care units of Duke University Hospital for collecting the blood samples used for this study and thank Dr. Patty Lee for supporting the ICU Biorepository. We thank Dr. Tony Moody for access to the pre-pandemic negative control samples used in this study, Dr. Thomas Denny for providing access to BSL2+ laboratory facilities to run the pre-pandemic negative samples, and Heidi Register for assistance with testing the negative samples on the DA-D4 POCT.

## Author Contribution

JTH and DSK are co-lead authors, who equally participated in experimental design, data collection, data analysis, manuscript drafting, figure creation and manuscript revision. LBO participated in experimental design, and collection and analysis of data for clinical validation. JL developed the D4Scope used throughout the study and drafted text and figures related to the D4Scope. DSK, CMF, and AMH developed the microfluidic cassette. GK cloned, expressed, and purified the N-NTD. SAW participated in data collection and analysis. CMF, DYJ, and AMH were responsible for conceptualization, investigation, and manuscript revision. CFP oversaw statistical analysis and assisted in study design. TO designed and ran live virus microneutralization assays, analyzed data and participated in writing the manuscript. GDS participated in writing the manuscript. BDK, CWW, LC, LGQ, SKN, BAS, IAN, and LBO contributed to the development of the Duke COVID-19 biorepositories and oversaw clinical data acquisition. BDK and CWW participated in manuscript revision. AC is the principal investigator who directed the studies, helped plan experiments, analyzed data, and participated in writing and editing the manuscript. All authors read and approved the manuscript.

## Ethics declarations

Immucor Inc., has acquired the rights to the D4 assay on POEGMA brushes for in vitro diagnostics from Sentilus Inc. (cofounded by AC and AMH).

## Notes

### Author Declarations

De-identified heat-inactivated EDTA plasma samples were accessed from the Duke COVID-19 ICU biorepository (Pro00101196, PI Bryan Kraft) via an exempted protocol approved by the Duke University Institutional review board (Pro00105331, PI Ashutosh Chilkoti). The exempt protocol was used to test all other negative control pre-pandemic samples.

