## Supplementary information for "Multiplexed, quantitative serological profiling of COVID-19 from a drop of blood by a point-of-care test"

**Contents:**

1) DA-D4 layout **(Fig. S1)**
2) Expression and purification of N-terminal domain of nucleocapsid protein (N-NTD) **(Methods, Fig. S2)**
3) Microfluidic chip fabrication **(Methods, Fig. S3)**
4) D4Scope fabrication (**Methods,** **Fig. S4)**
5) Patient profile for clinical validation study **(Table S1, Table S2)**
6) Representative images of DA-D4 array for a positive and negative sample (**Fig. S5)**
7) Microfluidic flow cell operation with whole blood **(Methods, Fig. S6)**
8) Correlation of DA-D4 readout with microneutralization assay **(Fig. S7)**

### 1) DA-D4 layout

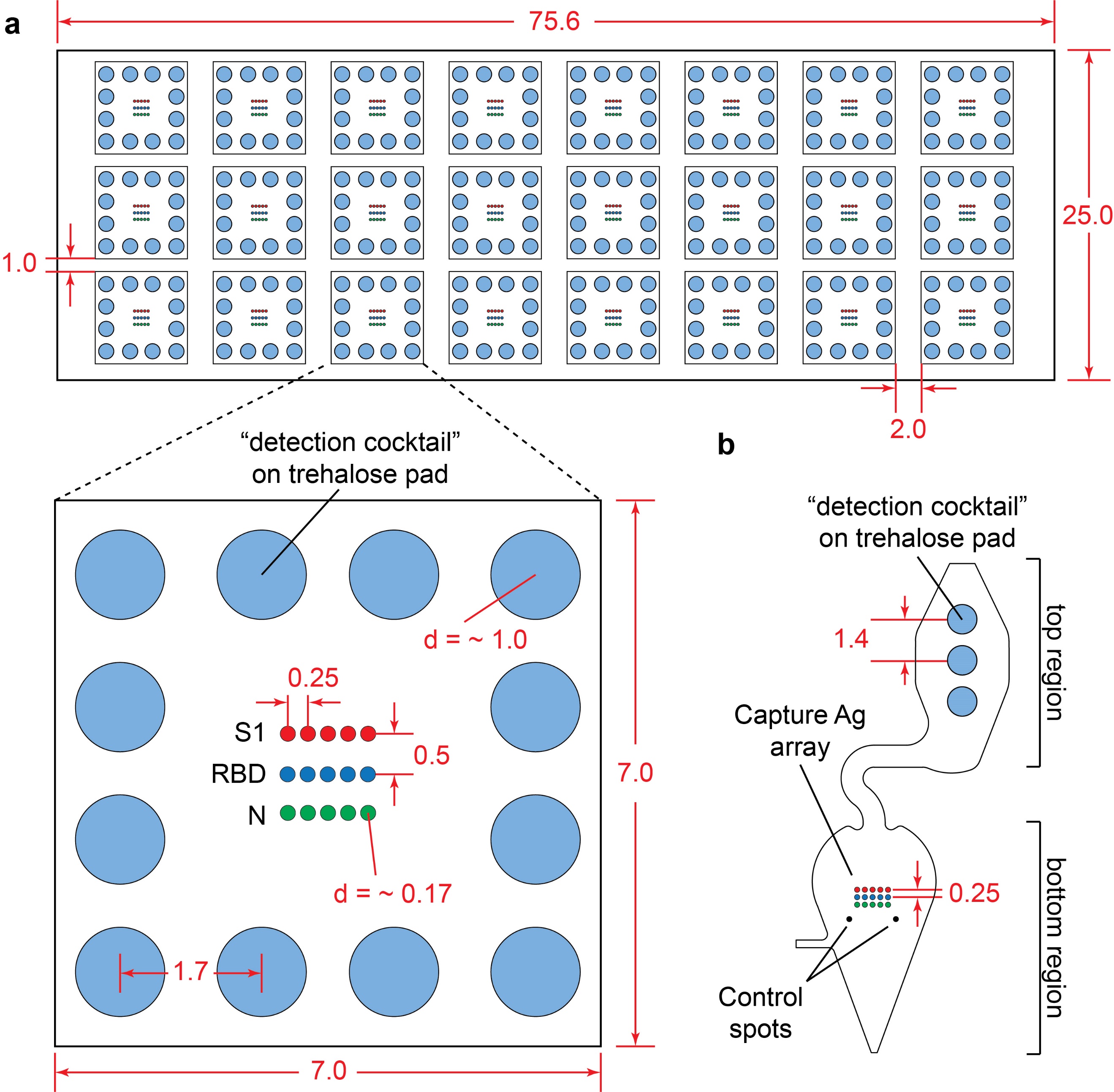

**Fig. S1.** **DA-D4 print layout in open format and microfluidic flow cell.** **(a)** open format print layout with zoomed in view of individual arrays. For assays with IP-10, an additional row of five capture antibody spots was printed and IP-10 detection antibody was added to the detection cocktail. **(b)** Microfluidic chip print format. Three passes of trehalose pads were printed, followed by eight passes of the detection cocktail. All measurements listed are in mm.

### 2) Expression and purification of N-terminal domain of nucleocapsid protein

*Methods:* The nucleotide sequence for the N-terminal domain of the nucleocapsid protein (N-NTD) of SARS-CoV-2 (residues 33-212) with a C-terminal His-tag was codon-optimized for E. coli. The gene was synthesized and cloned into a PET-24a vector (Twist Bioscience) and expressed in BL21(DE3) E. coli. Cultures were grown in shaker flasks at 25 ⁰C for 4 h, induced with 0.5 mM IPTG and grown overnight at 16 ⁰C. Cells were harvested and lysed by sonication, lysates were clarified by centrifugation, and N-NTD was purified from the lysate supernatant by immobilized metal affinity chromatography (IMAC) (**Fig. S2**).

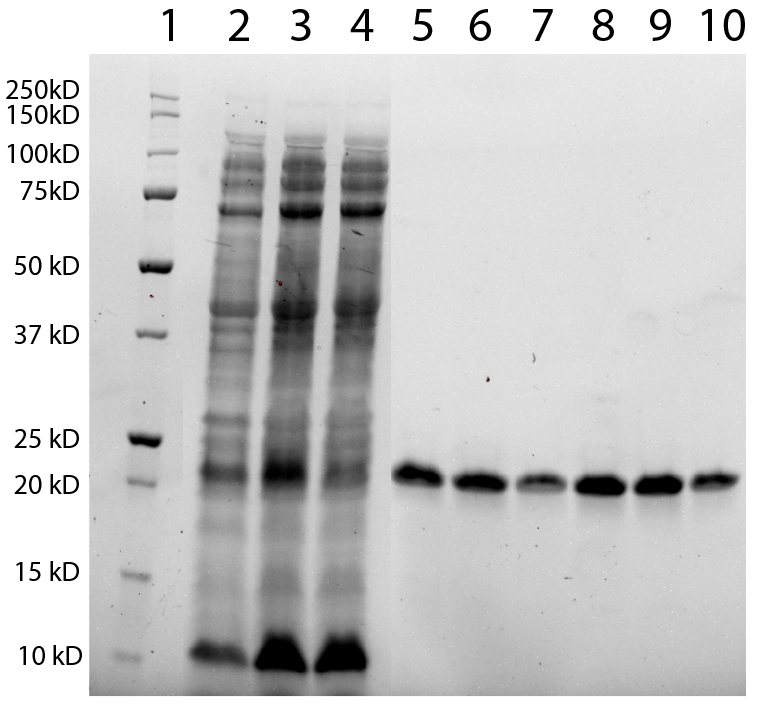

**Fig. S2.** **N-NTD purification.** The N-terminal domain of the SARS-CoV-2 nucleocapsid protein (N-NTD) was expressed in *E. Coli* and purified by IMAC, and aliquots from each step of the purification were visualized by SDS-PAGE. Lanes: ladder (lane 1), cell lysate (lane 2), clarified lysate (lane 3), IMAC flow through (lane 4), IMAC elution fractions (lanes 5-10).

### 3) Microfluidic flow cell fabrication

*Methods:* The microfluidic cassettes are fabricated from 1mm CLAREX acrylic sheets (Astra Products), 9474LE double-sided adhesive tape (3M company) and Whatman CF7 100 % cotton absorbent liner (Cytiva Life Sciences). 2D DXF files were generated in AutoCAD 2020 (Autodesk, Inc.) and laser-cut using a Gravostyle 8.0 and a LS900 Gravograph CO_2_ laser (Gravotech, Inc). 3D printed reservoirs and alignment tools were designed in SolidWorks 2019 (Dassault Systèmes SE) and printed on a Form 3 SLA 3D printer (Form Labs, Inc.).

Capture and detection reagents were printed onto the POEGMA coated glass slide in the same way as described in “*DA-D4 assay*” of the main text, with the only difference being the relative placement of the reagents. For the microfluidic flow cell, the detection reagents were printed in the top region of the reaction chamber and the capture spots were printed in the bottom region of the reaction chamber, as shown in **Fig. S1b**. This placement is a result of the reaction in the flow cell not being purely diffusion-driven, as is the case with the open format. Here, the combination of gravity and a downward direction of fluid flow requires upstream placement of the detection reagent to achieve even and efficient delivery of the detection reagents to the capture spots. The offset nature of the reaction chamber addresses an issue where small amounts of detection reagent that are not completely released from the POEGMA brush travel downstream in the brush during incubation and can lead to a higher background in the imaging area.

The microfluidic fluid cell assembly consists of five unique stacked layers with two additional attachments (wash reservoir and absorbent pad) shown in the exploded view of **Figure S3a**. (i) The fouling-resistant POEGMA coated glass is the base substrate for the flow cell, where reagents are printed, and serves as the back wall of the microfluidic channels. (ii) Adhesive #1 contains the pattern for the 400 µm wide microfluidic channels (timing channel, sample inlet, channel between wash chamber and reaction chamber). The thickness of the adhesive gives the channels their 157 µm depth. The outline of the reaction chamber and wash reservoir are left exposed to allow the sample and wash buffer to contact the POEGMA substrate. There is an open area at the outlet of the timing channel where the absorbent pad is adhered. The opening is slightly smaller than the pad,s creating a frame for the pads to be adhered to. (iii) Acrylic #1 provides the front wall of the microfluidic channels enclosing them on all four sides. The reaction chamber is left exposed at this layer to increase the reaction chamber volume to ~60 µL by adding the 1 mm of depth from the acrylic. This ensures that most of the sample remains in the reaction chamber during incubation as some sample traverses the timing channel. Additionally, this layer has cut outs for the sample inlet, wash reservoir, and absorbent pad that all need to be accessible to either complete assembly or operate the cassette. (iv) Adhesive #2 is a smaller layer that primarily serves as a seal between the two acrylic layers. It has a small crescent shaped flap at the sample inlet to create a one-way valve that prevents backflow after sample addition. The outline of the reaction chamber ensures there is an optically transparent path for the excitation laser to travel through during imaging preventing scattering of laser light on the less transparent adhesive that could impact performance. The outline of the wash reservoir provides access to the wash inlet needed for cassette operation. (v) Acrylic #2 seals the reaction chamber and features the sample inlet and wash reservoir access. (vi) A 3D-printed wash reservoir capable of holding up to 250 µL of wash buffer is attached to the exposed wash inlet (vii) Laser-cut absorbent pads absorb waste from the cassette and is held in place with an acrylic wash cover to reduce exposure of the waste to the end user.

**Figure S3b** provides a simplified overview of how the layers of the microfluidic flow cell were fabricated and assembled. (i) Adhesive layers #1 and #2 were fabricated by affixing an adhesive sheet, with its protective liners, to a rigid substrate using a sacrificial layer of double-sided adhesive. This assembly was then laser-cut with the features shown in **Figure S3a**. The rigid backing maintains the integrity of the otherwise flimsy microfluidic features. (ii) Acrylic layers #1 and #2 were laser cut separately without any special considerations. All features were rinsed with a 70% IPA solution and dried with nitrogen gas before assembly. (iii) The protective liners present on the acrylic layers and the top of the adhesive layer were removed. (iv) Acrylic layers #1 and #2 were then affixed onto adhesive layers #1 and #2 respectively. The newly created complementary acrylic/adhesive sub-assemblies can then be easily released from the backing protective liner of the adhesive that is still attached to the rigid backbone (iv) A custom designed 3D-printed positioning tool was then used to align the two sub-assemblies to each other and with the POEGMA substrate. To complete the final assembly, the adhesive backed 3D-printed reservoir was attached at the top of the fluid cell where wash buffer is dispensed. Two absorbent pads are fixed to the outlet of the cassette using exposed adhesive from the Adhesive #1 layer. An adhesive backed acrylic cover was placed over the pads to contain waste. For storage, the assembly is packaged in a thermally sealed pouch with 1 g of silica desiccant, and stored at ambient temperature and humidity.

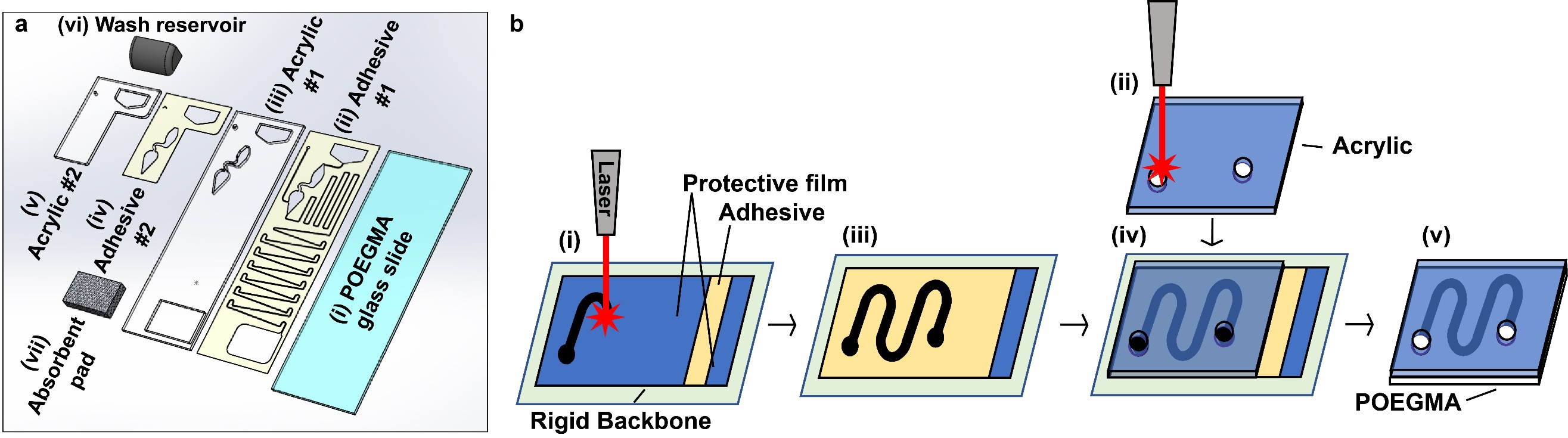

**Fig. S3. Schematic of microfluidic flow cell.** (a) Detailed exploded view of all layers and auxiliary attachments of the microfluidic flow cell (b) Simplified schematic detailing the process flow for the laser-cutting and assembly of the individual layers of the microfluidic flow cell.

### 4) D4Scope fabrication

*Methods:* The D4Scope is constructed using a Basler Ace CMOS Camera module AcA3088-57um (Basler AG), 676/37-25 nm bandpass filter (Semrock), MC100X lens (Optoengineering), 185mW 638nm red laser module (Sharp), Raspberry Pi 4B 2GB (Raspberry Pi Foundation), 3.5” TFT LCD display (UCTRONICS), and custom 3D printed housing parts made of polylactic acid filament (HATCHBOX PLA). The D4Scope is designed using SolidWorks (Dassault Systèmes SE) in assembly mode. The exploded view is shown in **Fig. S4** below. First, the optical components are positioned by mates to mount the objective lens at the listed working distance (47 mm) away from the D4 chip, the optical filter in-line and concentric with the lens, and the red laser obliquely angled 30° from the imaging axis. Then, the housing parts are designed around these optical components. Special considerations are made to ensure parts are (1) modular for quick prototyping, (2) 3D printable with minimal supports and post-processing, (3) reasonably assembled. The 3D printed parts are printed on a CR-10 Mini (Creality) and Taz 6 (Lulzbot) fused deposition modeling 3D printers.

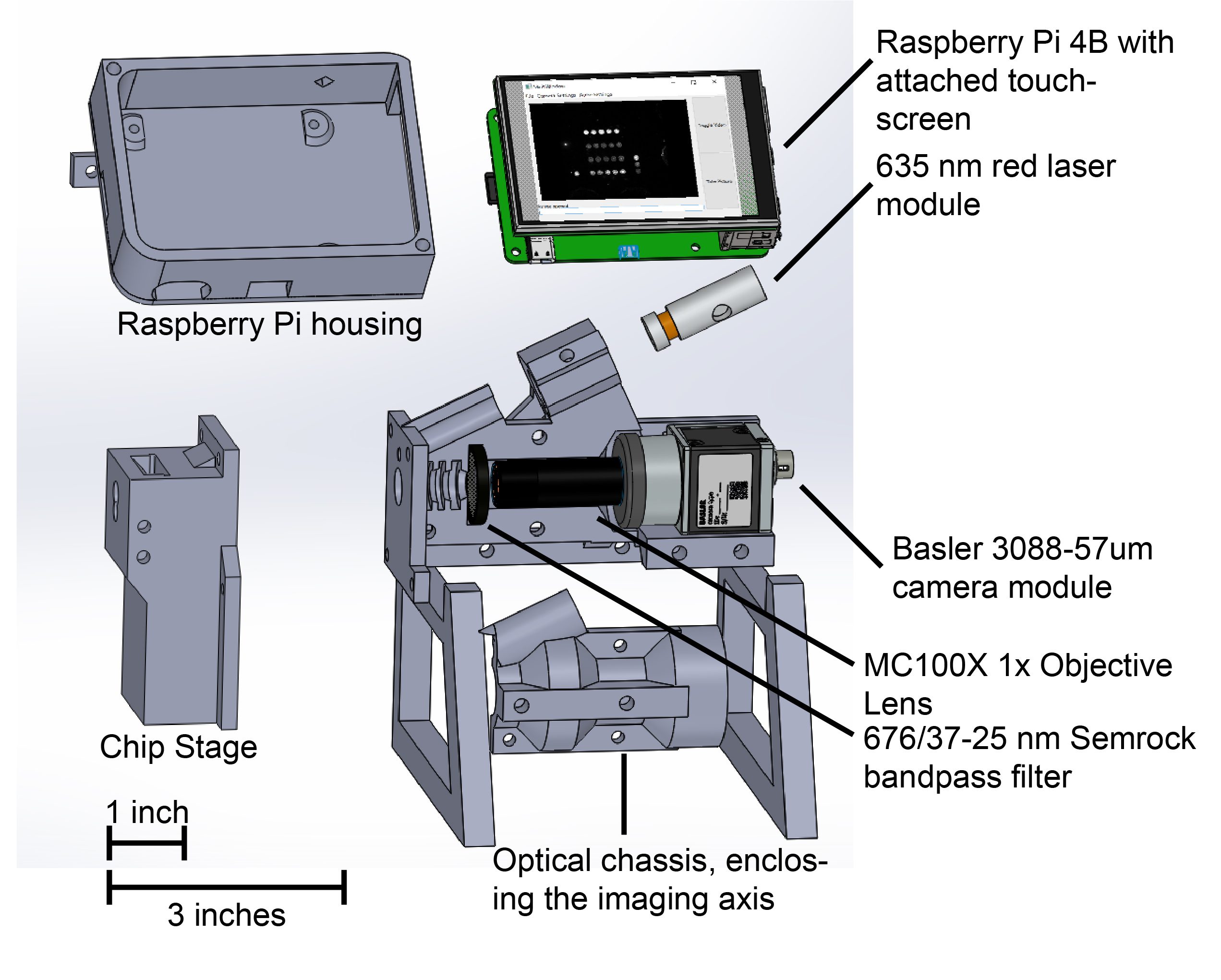

**Fig. S4. Exploded view of the D4Scope.** Files were rendered by SolidsWorks (Dassault Systems). All parts in grey are 3D printed in black poly-lactic acid plastic. M3 screws are used to fix all components in place: holding the D4 chip in focus relative to the camera/lens, enclosing the imaging axis (the path between the chip and the lens), and mounting the chip stage and Raspberry Pi (RPi) housing in place. M2 screws are used to mount the RPi to the housing.

### 5) Patient profile for clinical validation study

#### Table S1. COVID-19 patient clinical characteristics (Fig. 2)

| Characteristic | Patients (N=19) |
| --- | --- |
| Median age (range) | 55 (39-78) |
| Male sex | 9 |
| Race/ethnicity |  |
| Black | 13 |
| Latino | 3 |
| White | 2 |
| Other | 1 |
| Coexisting conditions |  |
| Any coexisting condition | 17 |
| Diabetes | 11 |
| Obesity | 12 |
| Hypertension | 7 |
| Cardiovascular disease | 6 |
| Renal Disease | 3 |
| Chronic Lung Disease | 3 |
| Stroke | 2 |

#### Table S2. Clinical characteristics of COVID-19 patients in blood testing study (Fig. 3)

| Characteristic | Patients (N=5) |
| --- | --- |
| Median age (range) | 56 (26-76) |
| Male sex | 4 |
| Race/ethnicity |  |
| Black | 1 |
| Latino | 0 |
| White | 3 |
| Other | 1 |

### 6) Representative images of DA-D4 array for a positive and negative sample

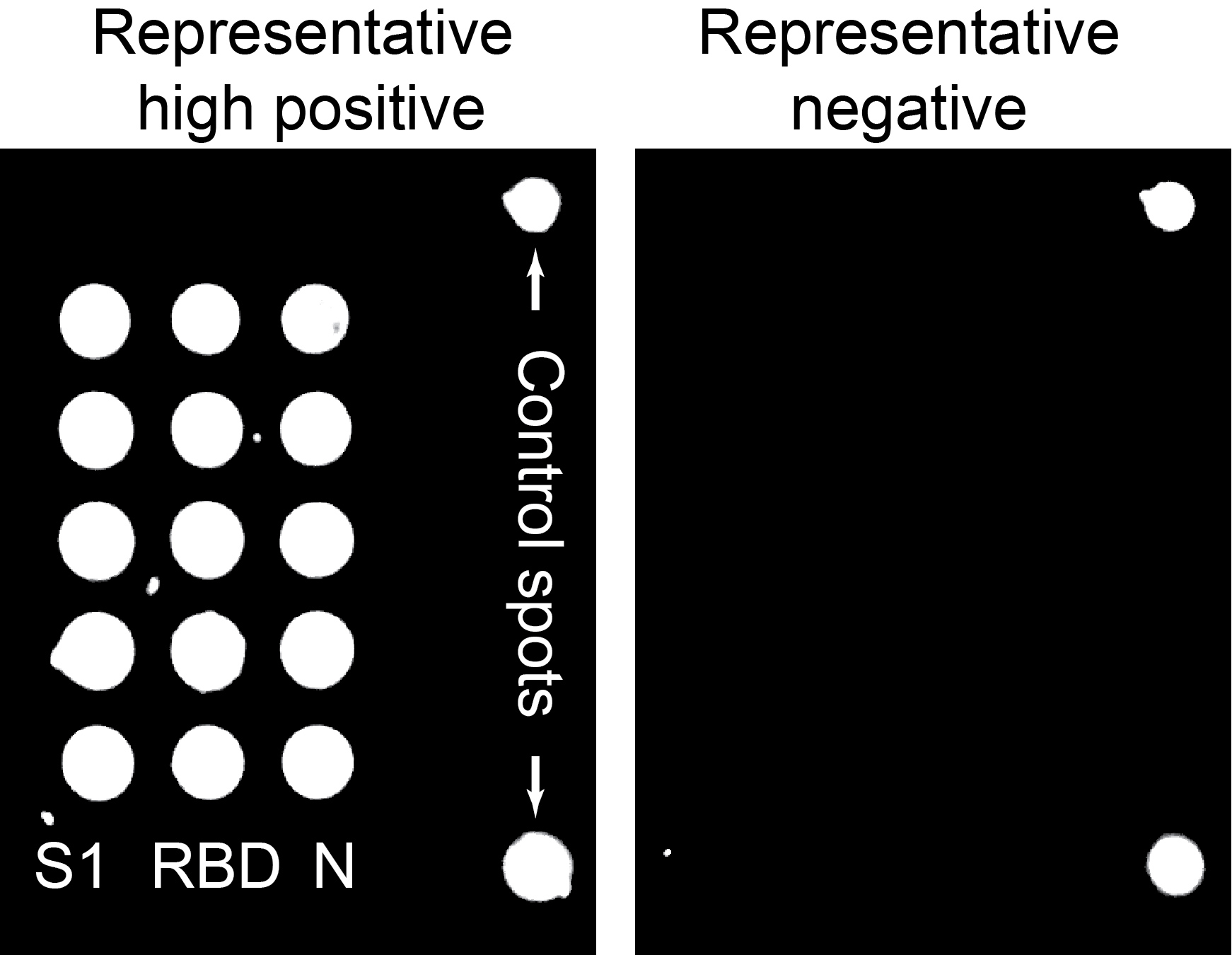

**Fig. S5. Representative high positive and negative arrays from patient samples.** (Left) DA-D4 array for a positive clinical sample. (Right) DA-D4 array for a negative clinical sample. The diameter of each capture antigen is ~170 µm. Adjustments to image brightness and contrast where performed the exact same for both images.

### 7) Microfluidic flow cell operation with whole blood

To test whole human blood from EDTA-collection tubes, three modifications were made to the microfluidic flow cell (**Fig. S6**). First, the timing channel was shortened and the large vertical loops were removed. The high viscosity of blood and tendency for red blood cells (RBCs) to pack near the capillary fluid front reduces flow rate and increases the likelihood of clogging when the direction of flow is against gravity. This change resulted in a reliable incubation time that is comparable to that observed with the plasma flow cell. Second, a 30° slope was added to the offset channel that separates the top region and bottom region of the reaction chamber. The original design, with no slope, collected RBCs that settled during incubation and were not removed during the wash step. These RBCs would only be flushed into the bottom region of the reaction chamber during the final drying step. This resulted in an undesirable increase in fluorescence background and decreased performance metrics of the assay. The 30° slope prevents this settling from occurring. Third, the timing channel outlet, which interfaces with the absorbent pad, was modified to prevent blockage of the channel by densely packed RBCs at the capillary fluid front. In the original design, when this dense cell layer reached the outlet-absorbent pad interface, wicking would not commence due lack of plasma at the interface. By approaching the wicking pad from the side, and adding an 80° slope to the outlet-wicking pad interface, the RBC dense fluid front can flow down the slope allowing more plasma-rich sample to reach the wicking pad interface and activate wicking.

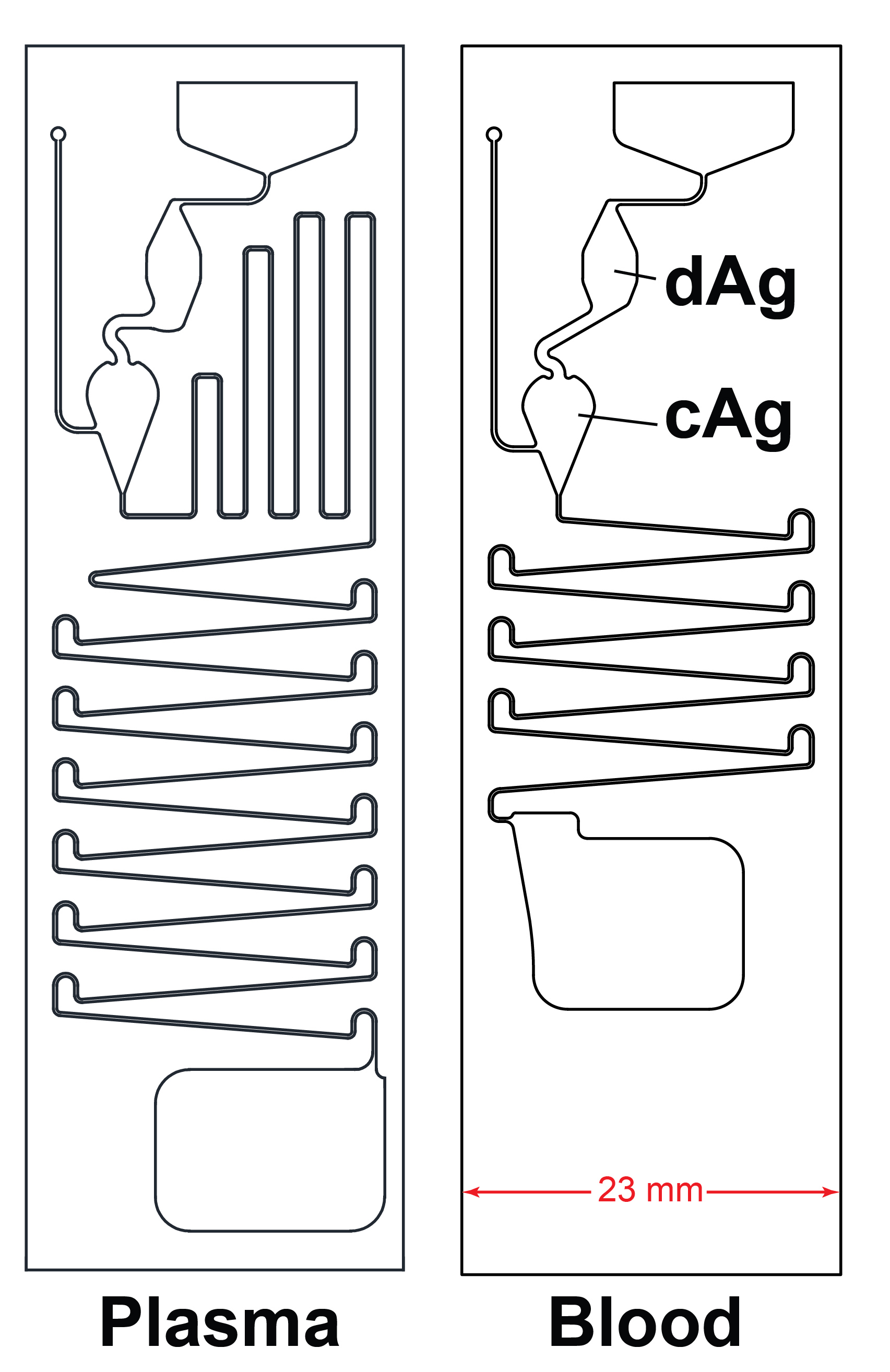

**Fig. S6. Differences in microfluidic flow cell for plasma/serum and blood.** CAD rendering of microfluidic flow cell used for testing plasma/serum (left) or blood (right).

### 8) Correlation of DA-D4 readout with microneutralization assay

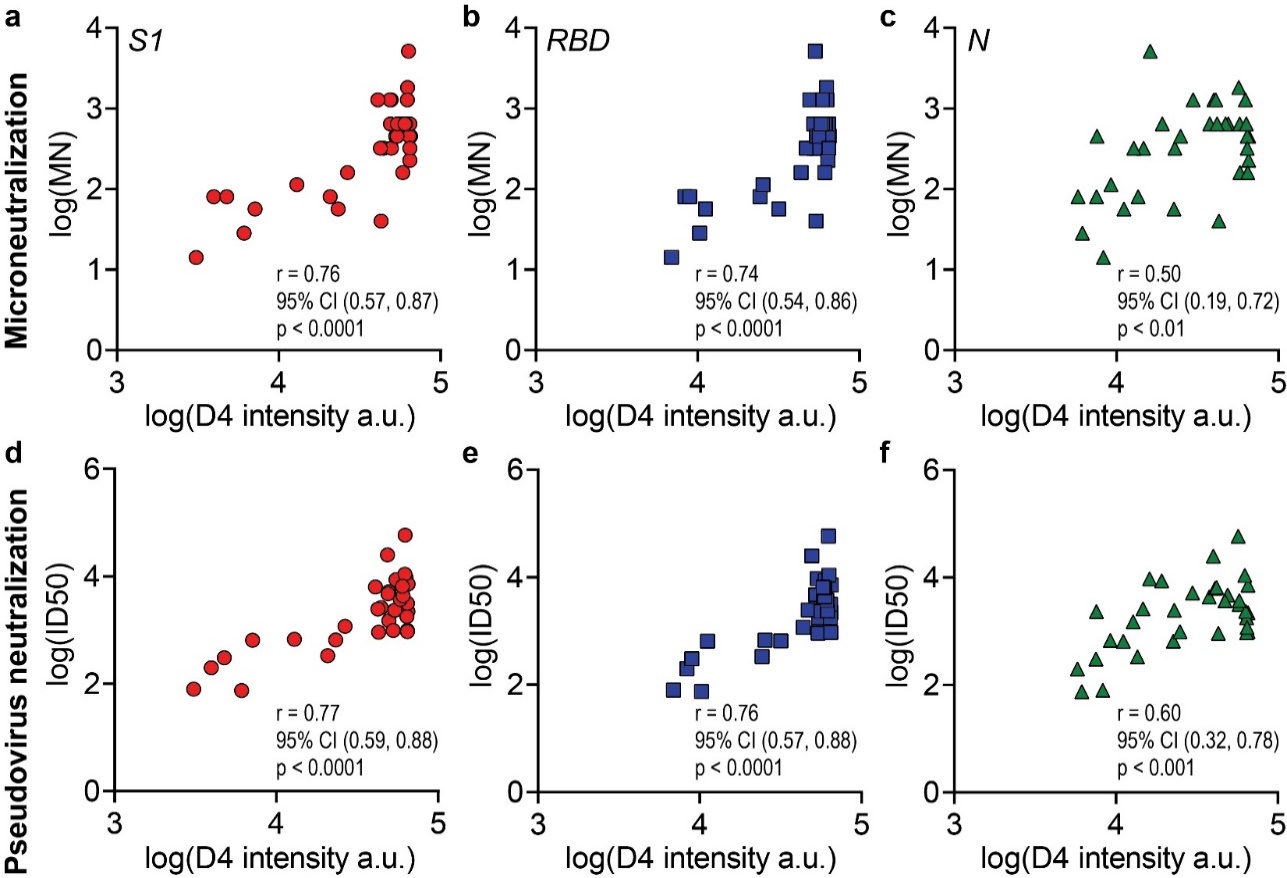

**Fig. S7. DA-D4 readout correlation with microneutralization assay.** Correlation between (**a**) anti-S1, (**b**) anti-RBD, and (**c**) anti-N with microneutralization assay for patient samples from Figure 2 with Pearson r reported.
